# Impact on alcohol selection and online purchasing of changing the proportion of available non-alcoholic versus alcoholic drinks: A randomised controlled trial

**DOI:** 10.1101/2022.03.04.22271898

**Authors:** Natasha Clarke, Anna KM Blackwell, Jennifer Ferrar, Katie De-Loyde, Mark A Pilling, Marcus R Munafò, Theresa M Marteau, Gareth J Hollands

## Abstract

**Background:** Increasing the availability of non-alcoholic options is a promising population-level intervention to reduce alcohol consumption, currently unassessed in naturalistic settings. This study in an online retail context aimed to estimate the impact of increasing the proportion of non-alcoholic (relative to alcoholic) drinks, on selection and purchasing of alcohol.

**Methods and Results:** Adults (n=737) residing in England and Wales who regularly purchased alcohol online were recruited between March-July 2021. Participants were randomly assigned to one of three groups: ‘25% non-alcoholic/75% alcoholic’; ‘50% non-alcoholic/50% alcoholic’; ‘75% non-alcoholic/25% alcoholic’, then selected drinks in a simulated online supermarket, before purchasing them in an actual online supermarket. The primary outcome was the number of alcohol units selected (with intention to purchase); secondary outcomes included actual purchasing. 607 participants (60% female, mean age = 38 years [range: 18-76]) completed the study and were included in the primary analysis. In the first part of a hurdle model, a greater proportion of participants in the ‘75% non-alcoholic’ group did not select any alcohol (13.1%) compared to the ‘25% non-alcoholic’ group (3.4%; 95% confidence interval [CI] - 2.09, -0.63; p < 0.001). There was no evidence of a difference between the ‘75% non-alcoholic’ and the ‘50% non-alcoholic’ (7.2%) groups (95% CI 0.10, 1.34; p = 0.022) or between the ‘50% non-alcoholic’ and the ‘25% non-alcoholic’ groups (95% CI -1.44, 0.17; p = 0.121). In the second part of a hurdle model in participants (559/607) selecting any drinks containing alcohol, the ‘75% non-alcoholic’ group selected fewer alcohol units compared to the ‘50% non-alcoholic’ (95% CI -0.44, -0.14; p < 0.001) and ‘25% non-alcoholic’ (95% CI - 0.54, -0.24; p < 0.001) groups, with no evidence of a difference between the ‘50% non-alcoholic’ and ‘25% non-alcoholic’ groups (95% CI -0.24, 0.05; p = 0.178). Overall, across all participants, 17.46 units (95% CI 15.24, 19.68) were selected in the ‘75% non-alcoholic’ group; 25.51 units (95% CI 22.60, 28.43) in the ‘50% non-alcoholic’ group; and 29.40 units (95% CI 26.39, 32.42) in the ‘25% non-alcoholic’ group. This corresponds to 8.1 fewer units (a 32% reduction) in the ‘75% non-alcoholic’ compared to the ‘50% non-alcoholic’ group, and 11.9 fewer alcohol units (41% reduction) compared to the ‘25% non-alcoholic’ group; 3.9 fewer units (13% reduction) were selected in the ‘50% non-alcoholic’ group than in the ‘25% non-alcoholic’ group.

For all other outcomes, alcohol selection and purchasing were consistently lowest in the ‘75% non-alcoholic’ group.

Study limitations include the setting not being entirely naturalistic due to using a simulated online supermarket as well as an actual online supermarket, and that there was substantial dropout between selection and purchasing.

**Conclusions:** This study provides evidence that substantially increasing the proportion of non-alcoholic drinks – from 25% to 50% or 75% - meaningfully reduces alcohol selection and purchasing. Further studies are warranted to assess whether these effects are realised in a range of real-world settings.

**Trial registration:** ISRCTN:<u>11004483</u>; OSF: https://osf.io/qfupw

**Author summary:** *Why was this study done?:* Excessive alcohol consumption contributes to the global burden of non-communicable diseases, including cancer, heart disease and stroke. Interventions that change physical and economic environments have the potential to reduce alcohol consumption. Interventions targeting physical environments include availability interventions that involve changing the proportion of healthier options that are available, relative to less healthy options. A previous online study found that increasing the availability of non-alcoholic compared to alcoholic drinks reduced the hypothetical selection of alcoholic drinks, but there is an absence of evidence from naturalistic settings.

*What did the researchers do and find?:* This study evaluated the impact of increasing the proportion of non-alcoholic (relative to alcoholic) drinks, on selection and actual purchasing of alcohol. In a randomised controlled trial, 737 participants were randomly assigned to one of three groups with varying proportions of alcoholic versus non-alcoholic drinks ‘25% non-alcoholic/75% alcoholic’; ‘50% non-alcoholic/50% alcoholic’; ‘75% non-alcoholic/25% alcoholic’). Participants selected drinks from 64 options in a simulated online supermarket which was designed to look and function similarly to an online supermarket. Participants were then required to immediately purchase the same drinks in an actual online supermarket. It was found that increasing the proportion of non-alcoholic drinks – from 25% to 50% or 75% – reduced the amount of alcohol selected and bought, in this online supermarket setting.

*What do these findings mean?:* This study provides evidence that increasing the proportion of non-alcoholic drinks could reduce alcohol selection and purchasing, highlighting the potential for availability interventions to reduce alcohol sales at population level. Further studies are warranted to assess whether these effects are realised in a range of real-world settings

## Introduction

Excessive alcohol consumption is one of four sets of modifiable behaviours - along with tobacco use, physical inactivity and unhealthy diet - that make a major contribution to the global burden of non-communicable diseases, including cancer, heart disease and stroke [1,2]. Given the influence of environmental cues upon consumption and related behaviours, interventions that change physical and economic environments in which these behaviours occur have the potential to reduce alcohol consumption [3]. Altering the availability of alcohol products has been identified as a particularly potent approach [4], but has typically been examined in relation to demographic, temporal or spatial restrictions (e.g., by age, opening hours, or number or density of retail outlets), and not in terms of changing the range of available products. One intervention of this kind, potentially scalable to population-level and currently untested, involves increasing the proportion of non-alcoholic (relative to alcoholic) drink options that are available to select, purchase, and ultimately consume [5]. This can be achieved by either making more non-alcoholic options available, removing some alcoholic options, or by doing both and so retaining the same overall number of options [6]; the latter is assessed in the current study.

The promise of so-called ‘availability’ interventions that change proportions of unhealthy (relative to healthier) products, is highlighted by an emerging evidence base in relation to food [5]. A Cochrane systematic review found that reducing the proportion of available food products of a certain type (e.g., unhealthy snacks) resulted in markedly reduced selection of those foods [7], although the included evidence was limited in both quality and quantity. More recent field trials also suggest that decreasing the proportion of higher energy or meat-based foods reduces their consumption [8–11]. In terms of alcohol products, there is an absence of evidence, with no eligible studies identified in the aforementioned Cochrane review [7] or in a recent search update [5]. In what is, to our knowledge, the only previous study that has examined the potential of such an availability intervention applied to alcohol, the proportion of participants selecting an alcoholic drink decreased from 74% when one-quarter of the available drinks were non-alcoholic, to 51% when three-quarters were non-alcoholic [12]. However, this study only measured hypothetical and mandatory selection of a single drink from a limited range of eight options. Studies using meaningful outcomes and conducted within more naturalistic contexts that include wider product ranges are necessary to inform the development and implementation of real-world interventions and policies.

There is clear interest in increasing the availability of non-alcoholic drink options, from the perspective of both consumers and policymakers. While the current market for alcohol-free beer, wine and spirits represents a 3.5% share of the global alcohol industry and is therefore relatively small, it is rapidly growing [13]. For example, low and no-alcohol beer currently accounts for 3% of the total beer market [14], but this is forecast to increase by nearly 13% per annum over the next 3 years and is the fastest growing drinks segment in the UK [15]. In 2021, the no/low alcohol market grew by 6% globally, and in the UK, sales of non-alcoholic beer increased by 7% [16].

In 2020, the UK Government made a commitment with the drinks industry to increase the availability of alcohol-free and low-alcohol products by 2025, although details on what this would involve have not been published [17]. Currently most consumers purchase no or low alcohol drinks infrequently, although increased availability of these products is associated with an increase in their sales [18] and reductions in grams of alcohol purchased [19,20]. Non-alcoholic alternatives to alcohol (i.e., alcohol-free drinks and soft drinks marketed to adults) still only represent a small proportion of the market, however, which combined with their recent increase in popularity, suggests there is substantial scope for increasing their availability.

The aim of the current study was to estimate the impact of increasing the proportion of non-alcoholic drink options relative to alcoholic drink options, on the number of alcohol units that are i) selected (with the intention to purchase) and ii) purchased. We hypothesised that increasing the availability of non-alcoholic alternatives to alcohol would reduce the number of alcohol units selected and purchased.

## Methods

The trial was prospectively registered (ISRCTN:<u>11004483</u>). In addition, both the study protocol (https://osf.io/qfupw) and a detailed statistical analysis plan (https://osf.io/4yuca) were pre-registered on the Open Science Framework (OSF). The study was approved by the Faculty of Life Sciences Research Ethics Committee at the University of Bristol (reference no: 116124). Trial reporting follows CONSORT 2010 guidelines.

### Study design

The study used a parallel-groups randomised controlled design. Individual participants were randomly allocated without stratification to one of three groups differing in the proportion (%) of non-alcoholic versus alcoholic drink options available for selection: Group 1: ‘25% non-alcoholic/75% alcoholic’; Group 2: ‘50% non-alcoholic/50% alcoholic’; Group 3: ‘75% non-alcoholic/25% alcoholic’.

### Setting

The study was conducted online using simulated and real online supermarkets. First, participants completed a simulated supermarket selection task hosted on the Qualtrics online survey platform (see https://osf.io/2cy7t for example task images). The simulated supermarket was designed to look and function as similarly to the actual online supermarket as was possible within Qualtrics. Drinks were displayed in rows of four drink options and participants could add these to their basket, which displayed a total price after the selection had been made. Following this, participants were required to purchase drinks in Tesco online supermarket (Tesco.com), the largest national supermarket in the UK.

### Participants

To be eligible for the study, participants had to be adults aged 18 years and over residing in England or Wales, who consumed beer or wine regularly (i.e., at least weekly), and purchased these drinks at least monthly from Tesco.com, with a minimum spend of £20. Participants had to be willing to complete a shop at Tesco.com following completion of the selection task, book a delivery or click-and-collect slot, and send proof of purchase (their receipt) to the research team. Similar proportions of males and females of a range of ages were recruited via Roots Research (https://rootsresearch.co.uk/), one of the largest research agencies in the UK, with a high-quality panel of over 350,000 participants. Recruitment occurred between March-July 2021.

#### Sample size

A previous online study compared the impact on drink selection of altering non-alcoholic vs alcoholic drink availability [12]. The proportion of participants selecting an alcoholic drink decreased from 74% when non-alcoholic drink availability was low (25% of drink options), to 61% when availability was medium (50% of drink options), and 51% when availability was high (75% of drink options) (i.e., a difference of 13% and 10%, respectively, between adjacent groups). However, only a single drink was selected in this online study and there was no intention to purchase the selected drinks (i.e., decisions were purely hypothetical). As such, to our knowledge there was no comparable evidence available from which to estimate effects of this intervention on selection or purchasing behaviour of multiple drink options. Available resources allowed recruitment of around 600 participants. As an illustrative calculation, assuming 15% attrition, a sample of 510 participants (170/group) was sufficient to detect an effect of d=0.3 for the primary outcome for a two-group t-test with alpha of 5% and 80% power. Using pre-testing data (~5/group), the conservative SD estimate was 12.1units (i.e., the maximum group variance observed), indicating that the sample size was sufficient to detect a difference of 3.7 alcohol units selected between groups.

### Randomisation and masking

Randomised assignment of participants was completed via the default algorithm in Qualtrics with a ratio of 1:1:1. Participants were unaware of their group assignment throughout the study. The research team were blinded to allocation until participants had completed the primary outcome; the statistician completing the analysis was blinded to the allocation.

### Intervention

All participants viewed a total range of 64 drink options. This comprised i) a range of beers, ciders, alcohol-free beer and cider alternatives, and soft drinks (32 options), and ii) a range of wines, alcohol-free wine alternatives, and soft drinks (32 options), modelled on the available range of products on Tesco.com. Initial scoping work found that Tesco.com proportions of alcoholic versus non-alcoholic options were roughly 25% non-alcoholic (360 options) and 75% alcoholic (1058 options). Alcohol-free beer, alcohol-free cider and alcohol-free wine options used in the task were matched as far as possible on brand and size characteristics with the alcohol options available online at Tesco.com. Additional alcoholic beer, cider and wine options were selected based on the leading brands of lager, ale, mild and stout, cider and wine [21–23] in Great Britain according to the number of consumers. Adult soft drinks were selected based on options that were commonly displayed next to alcoholic drinks in physical supermarkets and most likely to be consumed as a substitute for alcohol (meaning that children’s soft drinks, milk, and fruit juice were excluded). Participants viewed varying proportions of non-alcoholic and alcoholic drink options depending on their assignment: Group 1: ‘25% non-alcoholic/75% alcoholic’; Group 2: ‘50% non-alcoholic/50% alcoholic’; Group 3: ‘75% non-alcoholic/25% alcoholic’. The proportions used were based on previous food and alcohol studies [12,24]. Within each range of alcoholic drinks there were the same number of beer as wine options, and within each range of non-alcoholic drinks there were the same number of soft drinks as alcohol-free options. Participants were randomised to the order in which each subcategory (soft drinks; alcohol free; alcoholic) was presented within each of the beer and wine categories and the order of drinks within each subcategory was also randomised. Each drink option was displayed as an image, below which was a text description of the drink (identical to Tesco.com), the % alcohol by volume (ABV) for drinks containing alcohol, and its price.

Full details of the task, as well as the complete list of drinks are in the Supplementary Material S1. In the Typology of Interventions in Proximal Physical Micro‐Environments (TIPPME) [3], this is classified as an *‘Availability x Product’* intervention, while in a detailed conceptual framework specific to availability interventions [6], this is categorised as a ‘Relative Availability’ intervention.

### Procedure

Participants were initially provided with an information sheet, instructions, and a link to the study via email. Participants were told the study was investigating “Adult drink preferences in England and Wales” and were not made aware of the study aim. Participant instructions outlined the stages of the study in detail, i.e., that participants were required to select the drinks for their next shop from Tesco.com in a simulated online supermarket (Stage 1), then to immediately go to Tesco.com to book a delivery slot and add these drinks to their shopping basket (Stage 2), and finally to send their receipt to the study team on their delivery or collection day (Stage 3). Once they had started the study task, participants were again presented with this information and provided consent. Participants were randomised and in a simulated online supermarket environment replicating Tesco.com (within Qualtrics) they were shown the available drink selection. They chose all the drinks they wanted to purchase in their next online shop at Tesco.com. They were then shown their total drink selection and price, and given the opportunity to amend their selection before continuing. Participants then completed demographic and drinking behaviour measures.

After completing the simulated online supermarket task, participants were automatically sent an email detailing their selection. They were prompted to open this email and given further instructions for completing purchasing, alongside a direct link to Tesco.com. Participants placed their selected drinks in their Tesco.com shopping basket, along with any other items, booked their delivery or collection slot, and confirmed this within 48 hours. They were sent a reminder email on their delivery/collection day and requested to send an itemised receipt to the research team within 48 hours. Up to two follow-up reminders were sent, two and four days later. Purchases were recorded from receipts, including any additional drink purchases. Substitutions by the participant or by Tesco that were explained (e.g., not in stock) were marked as the original drink they attempted to purchase. Participants were debriefed via email and reimbursed £25(~$35) for their time taking part in the study (but not the drinks they purchased).

### Outcome measures

#### Primary outcome

The primary outcome was the number of alcohol units selected in the context of a stated intention to purchase. In the UK, a unit is a standard measure of pure alcohol in a drink with one unit equivalent to 10ml or 8g of pure alcohol (this is equivalent to 0.56 of a US standard drink [25]). Participants were aware when selecting drinks in the task that they were required to subsequently purchase the drinks chosen and send proof of this to the research team (otherwise they were not reimbursed). Units of alcohol were calculated for all drinks that were >0% ABV, i.e., alcoholic and ‘alcohol-free’ drinks (which were defined as containing more than 0% and up to 0.5% ABV). This outcome was pre-registered as the primary outcome as it was assessed in all participants who were exposed to the intervention, and measured within the same context, i.e., the simulated online supermarket.

#### Secondary and additional outcomes

Secondary outcomes were the number of alcoholic and non-alcoholic drinks selected, the number of alcohol units purchased, and the proportion (i.e., percentage) of total drinks selected and purchased that were alcoholic. Additional outcomes were the total number of drinks selected, and purchased, the number of alcoholic drinks purchased, and the number of non-alcoholic drinks purchased.

Selection outcomes were assessed from the simulated online supermarket task and purchasing outcomes were assessed from receipts after shops at Tesco.com were completed. Purchasing outcomes were calculated to include (i) additional drinks from study categories only (i.e., beer, cider, wine, and adult non-alcoholic drinks), and (ii) all additional drinks (i.e., all alcoholic and non-alcoholic drinks - excluding squash, juice, tea, coffee and children’s drinks).

#### Other measures

##### Demographic characteristics

Age, sex, and highest qualification attained (‘Higher Education or professional / vocational equivalents’, ‘A levels or vocational level 3 or equivalents’, ‘GCSE / O Level grade A*‐C or vocational level 2 or equivalents’, ‘Qualifications at level 1 and below’, ‘Other qualifications: level unknown’, or ‘No qualifications’). Qualifications classifications were based on UK definitions [26,27].

##### Household members

Number of adults (aged 18+) and of children (aged <18).

##### Drinking behaviour risk

Alcohol Use Disorders Identification Test (AUDIT) [28], a 10-item clinical screening measure for assessing risk associated with participants’ drinking behaviour (low risk drinking: score 0-7; medium/hazardous risk drinking: score 8-15; high/harmful risk drinking: score ≥16).

##### Baseline weekly unit consumption

Self-reported drinks consumed and purchased over the previous seven days, used to calculate the number of alcohol units as a continuous variable.

##### Free-text comments

Participants provided comments on the task, such as explaining their choice of drinks.

### Statistical analysis

Analyses were pre-registered in a detailed statistical analysis plan (https://osf.io/4yuca).

All participants who completed the selection task were included in the primary outcome analysis. Participants who failed to complete the selection task and those whose responses were flagged as incomplete or suspicious - e.g., those that forged data (i.e., submitted fake receipts) or selected an unrealistically large number (with a cut-off of >100 drinks) of drinks that were not purchased – were excluded (see Figure 1 for details by group). The criteria used to exclude data were not pre-registered but were defined and applied prior to data analysis, while researchers were unaware of group allocation. The data included participants that did not select any drinks, as this was still a valid choice. The distribution of the primary outcome was highly skewed and zero inflated, and therefore a hurdle model was used for analysis, fitting i) a binary logistic model (part 1) to the zero and non-zero outcomes and ii) a truncated negative binomial model (part 2) to just the positive values [29,30]. The model results for the positive values are therefore based only on participants who selected at least one drink containing alcohol (see Supplementary S2), with non-integer variables rounded to integer values before hurdle model analysis. The marginal effect estimates, with 95% CI, are presented in Table 4.

**Figure 1.**
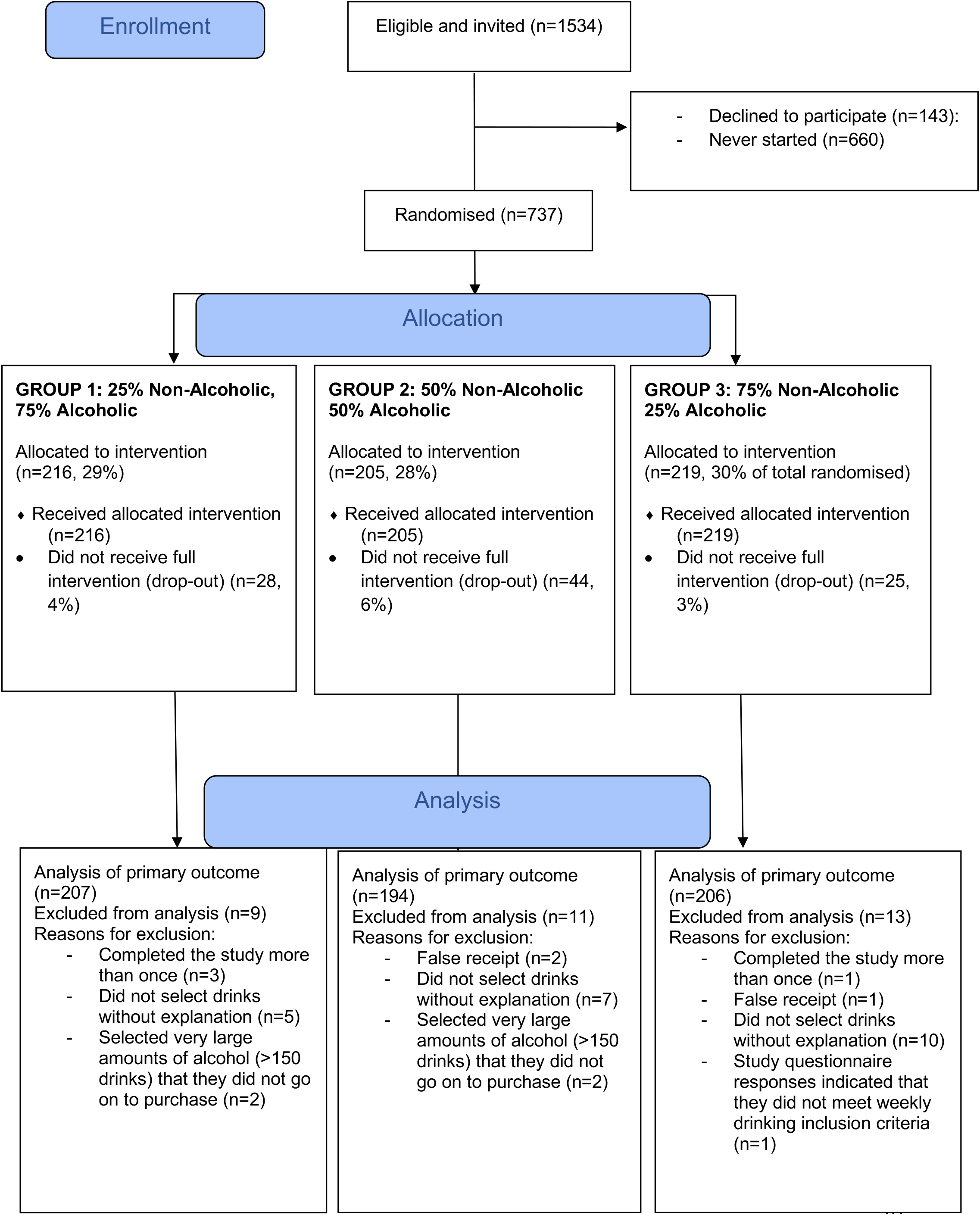
Participant flowchart.

For most secondary outcomes, hurdle models were repeated as per the primary outcome model. Model results for the binary outcomes (part 1 of the model) and the positive values (part 2, i.e., based on values above zero) are reported in Table 3. Marginal effect estimates for all secondary outcomes, with 95% CI, are presented in Table 4. The p-values for part 2 of the model and the change in marginal effect estimates (with the associated percentage reduction) are reported in the Results. For additional purchasing outcomes, negative binomial regression was required due to the skewed data. For the proportion outcomes (i.e., percentage of total drinks selected, and purchased, that were alcoholic), a beta binomial regression was used to model the proportion using the counts of relevant drinks selected out of the count of all drinks selected and this could accommodate the bimodal distribution observed. For these outcomes only, due to the nature of the model, any participants who did not select any drink (as appropriate for the outcome) were excluded.

Two per-protocol analyses were pre-specified, in which the primary outcome analysis was repeated for (i) participants who purchased what they selected, either with or without additional drinks (per-protocol analysis 1); (ii) participants who purchased exactly what they selected and purchased no additional drinks (per-protocol analysis 2).

For all outcomes, for the co-primary comparisons of primary interest (using the ‘25% non-alcoholic’ group as the reference group), a 5%/2 adjustment to the interpretation threshold for statistical significance was made. For the third comparison of tertiary interest (where ‘75% non-alcoholic’ and ‘50% non-alcoholic’ groups were compared), a simplistic 5%/3 adjustment was made rather than using methods which may not report all p-values (e.g., Benjamini-Hochberg, Holm-Bonferroni). These additional tests were calculated by refitting the same model with different reference categories.

## Results

### Sample characteristics

Figure 1 shows the flow of participants. In total, 737 participants were randomised, 640 of whom completed the selection task. 607 participants were included in the primary outcome analysis. The primary analysis dataset was 59.7% female and the mean age was 37.8 years (SD = 11.4; range: 18-76). Groups were well balanced on all characteristics (Table 1).

**Table 1.**
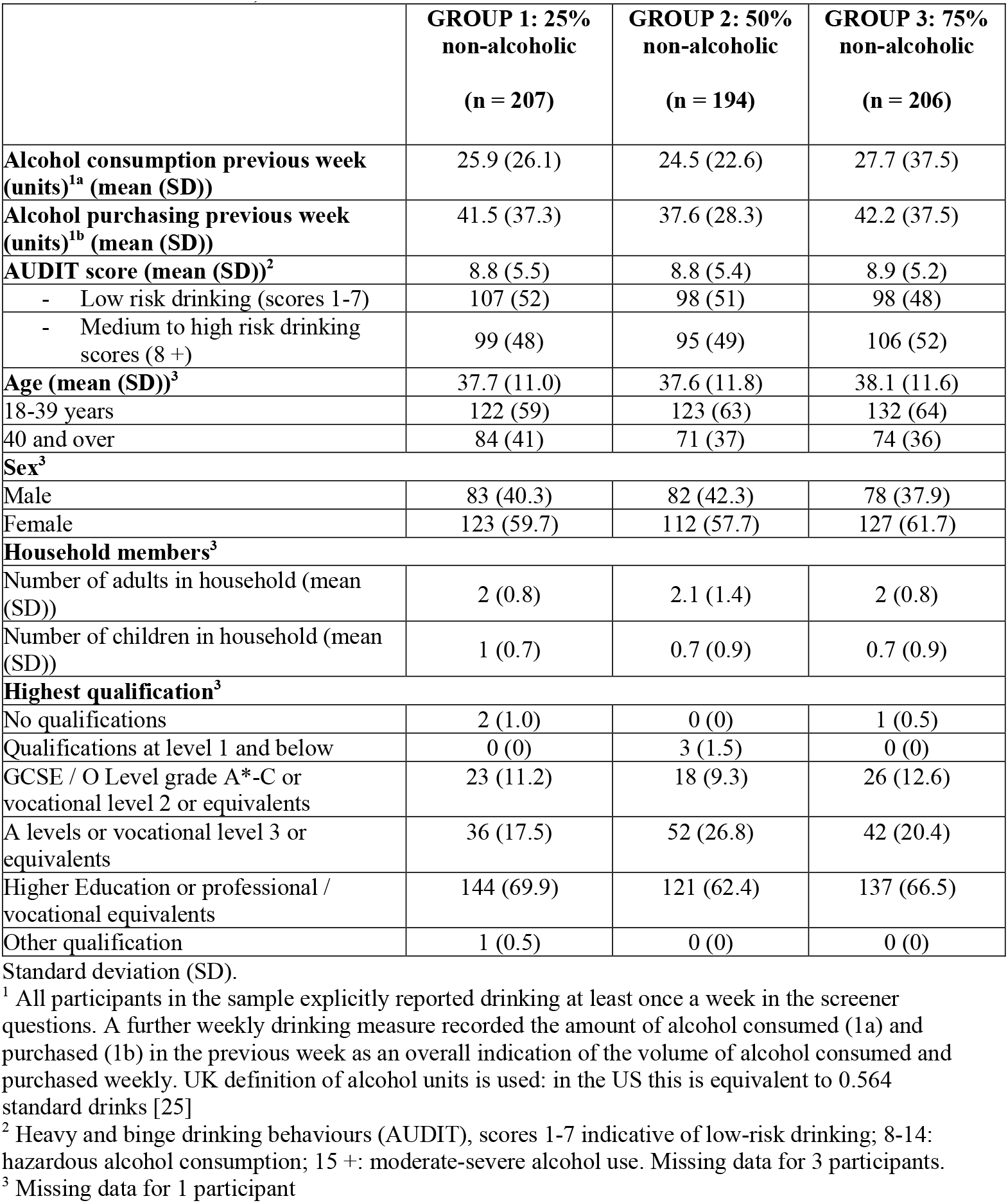
Characteristics of participants included in primary outcome analysis (n (%), unless otherwise stated)

|  | **GROUP 1: 25%**<br>**non-alcoholic**<br>**(n = 207)** | **GROUP 2: 50%**<br>**non-alcoholic**<br>**(n = 194)** | **GROUP 3: 75%**<br>**non-alcoholic**<br>**(n = 206)** |
| --- | --- | --- | --- |
| **Alcohol consumption previous week (units)<sup>1a</sup> (mean (SD))** | 25.9 (26.1) | 24.5 (22.6) | 27.7 (37.5) |
| **Alcohol purchasing previous week (units)<sup>1b</sup> (mean (SD))** | 41.5 (37.3) | 37.6 (28.3) | 42.2 (37.5) |
| **AUDIT score (mean (SD))<sup>2</sup>** | 8.8 (5.5) | 8.8 (5.4) | 8.9 (5.2) |
| - Low risk drinking (scores 1-7) | 107 (52) | 98 (51) | 98 (48) |
| - Medium to high risk drinking scores (8 +) | 99 (48) | 95 (49) | 106 (52) |
| **Age (mean (SD))<sup>3</sup>** | 37.7 (11.0) | 37.6 (11.8) | 38.1 (11.6) |
| 18-39 years | 122 (59) | 123 (63) | 132 (64) |
| 40 and over | 84 (41) | 71 (37) | 74 (36) |
| **Sex<sup>3</sup>** |  |  |  |
| Male | 83 (40.3) | 82 (42.3) | 78 (37.9) |
| Female | 123 (59.7) | 112 (57.7) | 127 (61.7) |
| **Household members<sup>3</sup>** |  |  |  |
| Number of adults in household (mean (SD)) | 2 (0.8) | 2.1 (1.4) | 2 (0.8) |
| Number of children in household (mean (SD)) | 1 (0.7) | 0.7 (0.9) | 0.7 (0.9) |
| **Highest qualification<sup>3</sup>** |  |  |  |
| No qualifications | 2 (1.0) | 0 (0) | 1 (0.5) |
| Qualifications at level 1 and below | 0 (0) | 3 (1.5) | 0 (0) |
| GCSE / O Level grade A*-C or vocational level 2 or equivalents | 23 (11.2) | 18 (9.3) | 26 (12.6) |
| A levels or vocational level 3 or equivalents | 36 (17.5) | 52 (26.8) | 42 (20.4) |
| Higher Education or professional / vocational equivalents | 144 (69.9) | 121 (62.4) | 137 (66.5) |
| Other qualification | 1 (0.5) | 0 (0) | 0 (0) |

Standard deviation (SD).
<sup>1</sup> All participants in the sample explicitly reported drinking at least once a week in the screener questions. A further weekly drinking measure recorded the amount of alcohol consumed (1a) and purchased (1b) in the previous week as an overall indication of the volume of alcohol consumed and purchased weekly. UK definition of alcohol units is used: in the US this is equivalent to 0.564 standard drinks [25]
<sup>2</sup> Heavy and binge drinking behaviours (AUDIT), scores 1-7 indicative of low-risk drinking; 8-14: hazardous alcohol consumption; 15 +: moderate-severe alcohol use. Missing data for 3 participants.
<sup>3</sup> Missing data for 1 participant

**Table 2.** Primary and secondary outcomes (raw means (SD))

|  | **GROUP 1: 25% non-alcoholic**<br>(n = 207) | **GROUP 2: 50% non-alcoholic**<br>(n = 194) | **GROUP 3: 75% non-alcoholic**<br>(n = 206) |
| --- | --- | --- | --- |
|  | **Mean (SD)** |  |  |
| **Primary outcome:** Number of alcohol units<sup>1</sup> selected (with an intention to purchase). | 29.5 (29.8) | 25.6 (20.5) | 17.6 (16.2) |
| **Secondary outcomes: selection** |  |  |  |
| Number of alcoholic drinks selected | 10.6 (14.0) | 8.8 (9.2) | 6.4 (7.1) |
| Number of non-alcoholic drinks selected | 5.4 (13.6) | 6.4 (10.5) | 8.8 (15.0) |
| Percentage of total drinks selected that are alcoholic | 75% (34%) | 64% (34%) | 52% (37%) |
| **Secondary outcomes: purchasing** |  |  |  |
|  | **GROUP 1: 25% non-alcoholic**<br>(n = 145) | **GROUP 2: 50% non-alcoholic**<br>(n = 141) | **GROUP 3: 75% non-alcoholic**<br>(n = 136) |
| Number of alcohol units purchased (including additional drinks from study categories only) | 26.7 (18.6) | 28.7 (23.3) | 23.4 (30.4) |
| Number of alcohol units purchased (including all additional drinks) | 29.1 (22.5) | 30.7 (26.9) | 28.7 (36.6) |
| Percentage of total drinks purchased that are alcoholic (including additional drinks from study categories only) | 76% (34%) | 68% (32%) | 55% (37%) |

<sup>1</sup> N.B. UK definition of alcohol units is used: in the US this is equivalent to 0.564 standard drinks [25]

|  |  |  |  |
| --- | --- | --- | --- |
| Percentage of total drinks purchased that are alcoholic<br>(including all additional drinks) | 68% (36%) | 61% (33%) | 52% (36%) |

**Table 3.**
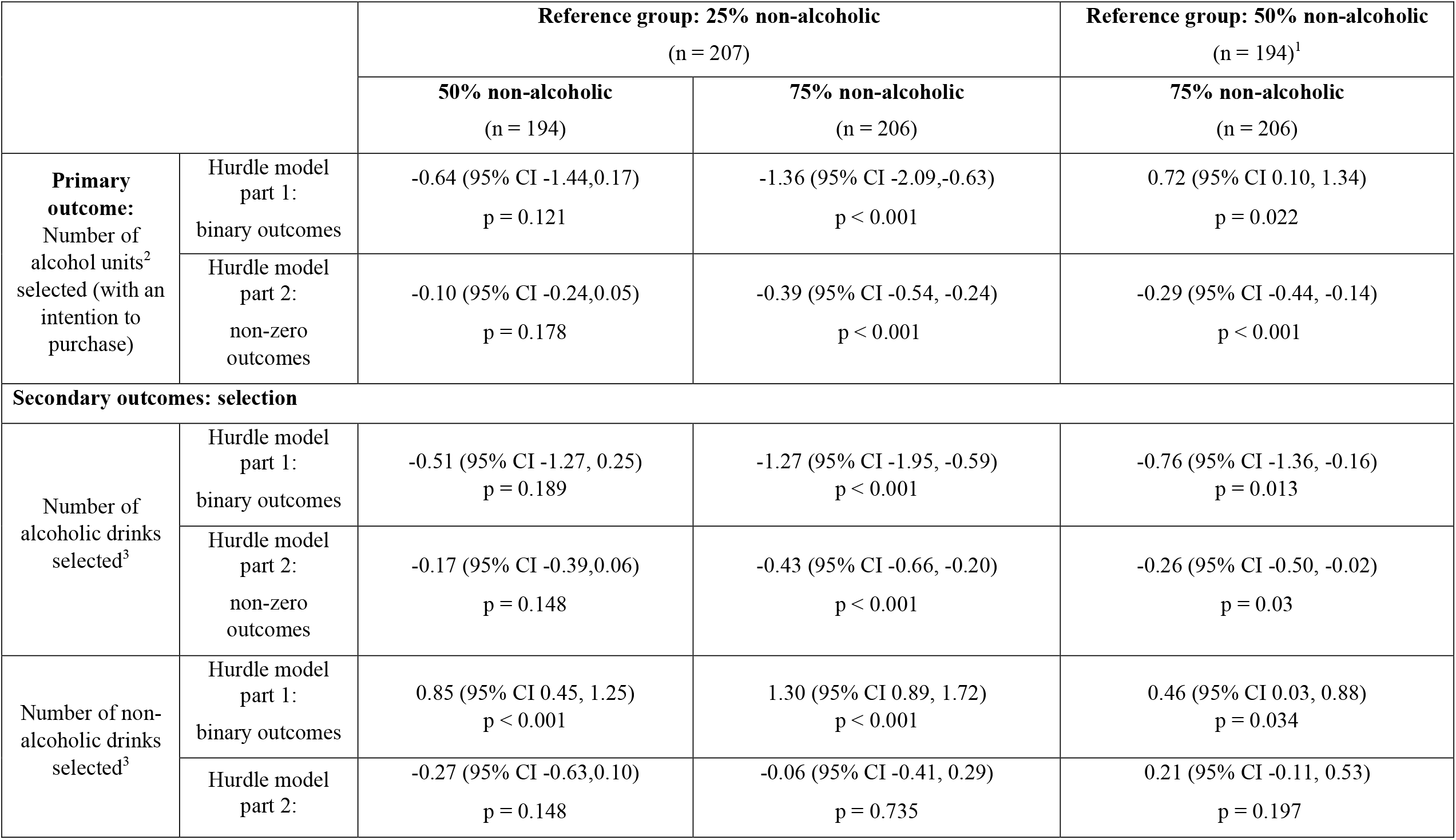
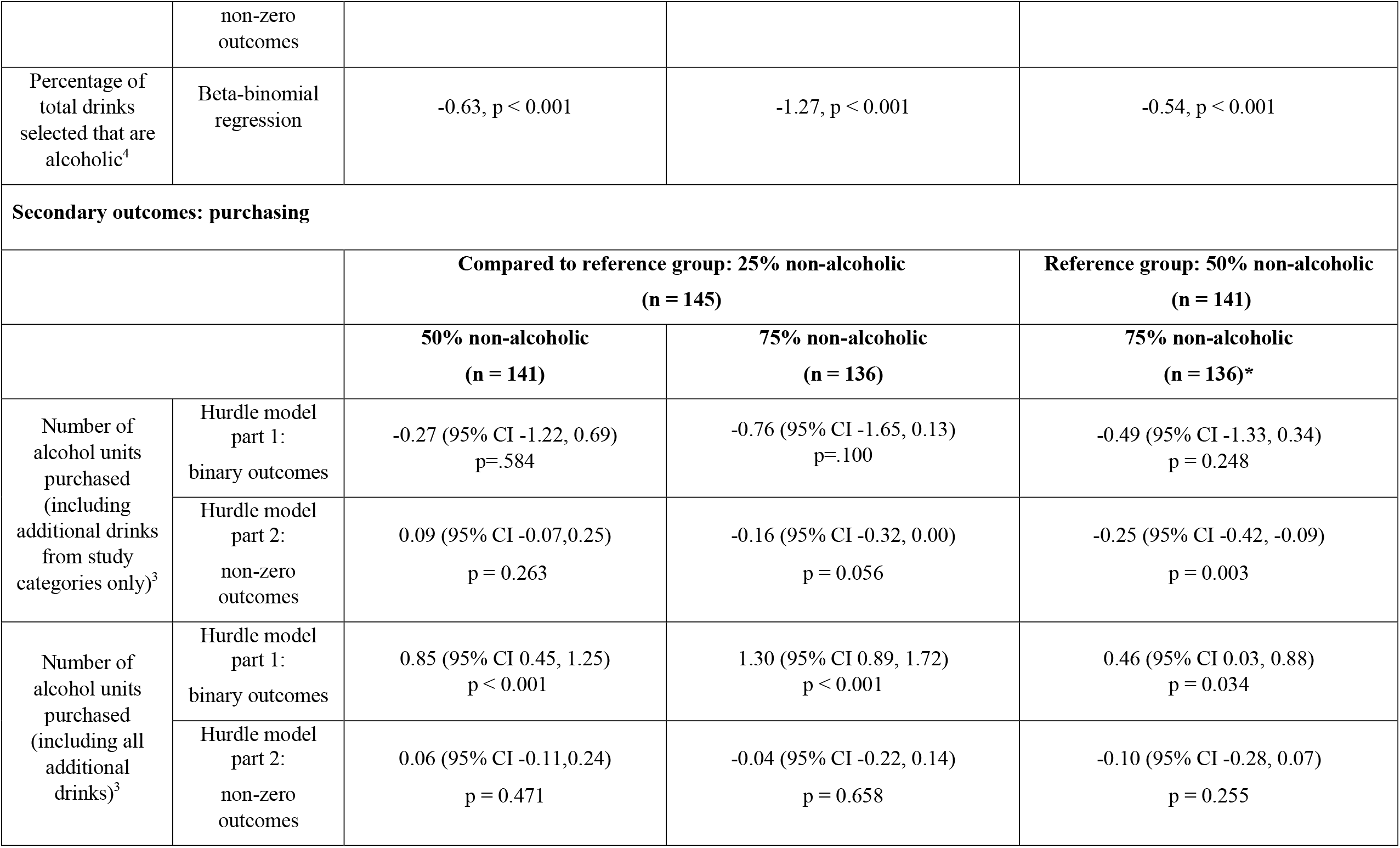
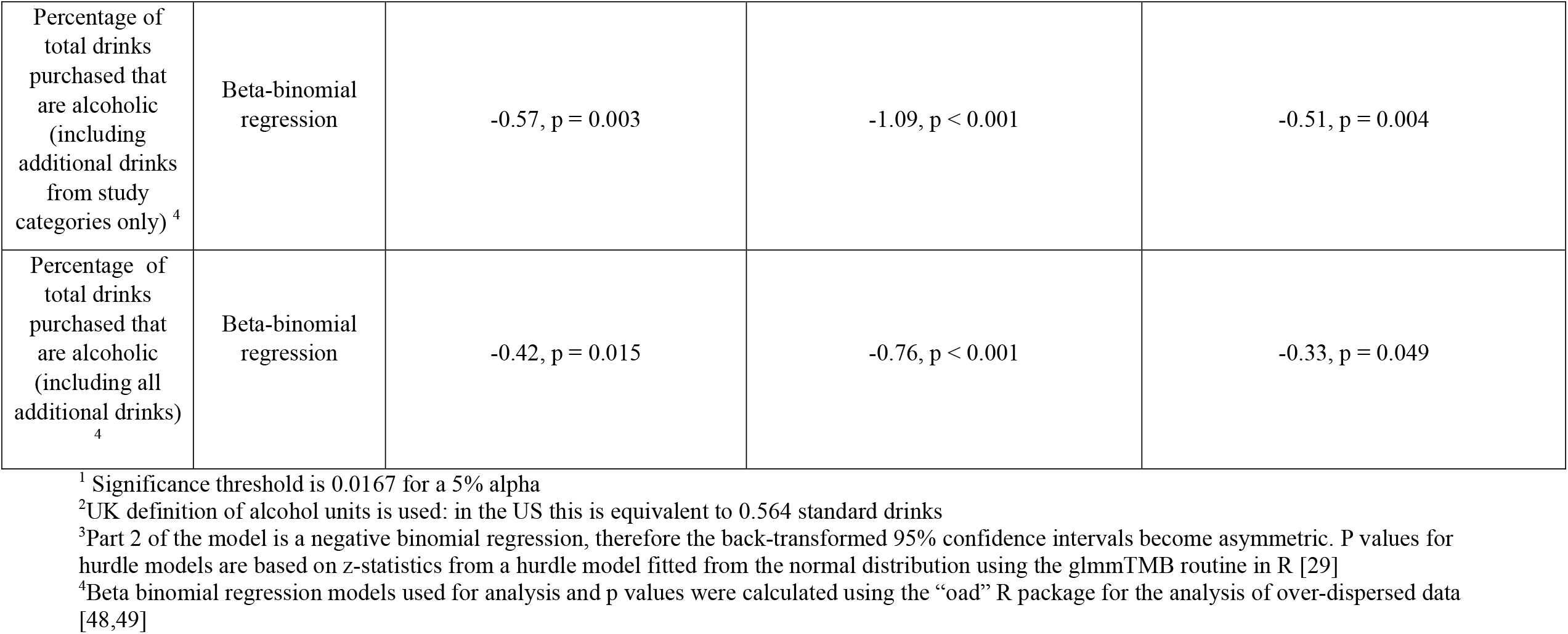
Model results for primary and secondary outcomes: estimates from hurdle models (95% confidence intervals), p values.

|  |  | Reference group: 25% non-alcoholic<br>(n = 207) |  | Reference group: 50% non-alcoholic<br>(n = 194)<sup>1</sup> |
| --- | --- | --- | --- | --- |
|  |  | 50% non-alcoholic<br>(n = 194) | 75% non-alcoholic<br>(n = 206) | 75% non-alcoholic<br>(n = 206) |
| **Primary outcome:**<br>Number of alcohol units<sup>2</sup> selected (with an intention to purchase) | Hurdle model part 1:<br>binary outcomes | -0.64 (95% CI -1.44,0.17)<br>p = 0.121 | -1.36 (95% CI -2.09,-0.63)<br>p < 0.001 | 0.72 (95% CI 0.10, 1.34)<br>p = 0.022 |
|  | Hurdle model part 2:<br>non-zero outcomes | -0.10 (95% CI -0.24,0.05)<br>p = 0.178 | -0.39 (95% CI -0.54, -0.24)<br>p < 0.001 | -0.29 (95% CI -0.44, -0.14)<br>p < 0.001 |
| **Secondary outcomes: selection** |  |  |  |  |
| Number of alcoholic drinks selected<sup>3</sup> | Hurdle model part 1:<br>binary outcomes | -0.51 (95% CI -1.27, 0.25)<br>p = 0.189 | -1.27 (95% CI -1.95, -0.59)<br>p < 0.001 | -0.76 (95% CI -1.36, -0.16)<br>p = 0.013 |
|  | Hurdle model part 2:<br>non-zero outcomes | -0.17 (95% CI -0.39,0.06)<br>p = 0.148 | -0.43 (95% CI -0.66, -0.20)<br>p < 0.001 | -0.26 (95% CI -0.50, -0.02)<br>p = 0.03 |
| Number of non-alcoholic drinks selected<sup>3</sup> | Hurdle model part 1:<br>binary outcomes | 0.85 (95% CI 0.45, 1.25)<br>p < 0.001 | 1.30 (95% CI 0.89, 1.72)<br>p < 0.001 | 0.46 (95% CI 0.03, 0.88)<br>p = 0.034 |
|  | Hurdle model part 2:<br>non-zero outcomes non-zero outcomes | -0.27 (95% CI -0.63,0.10)<br>p = 0.148 | -0.06 (95% CI -0.41, 0.29)<br>p = 0.735 | 0.21 (95% CI -0.11, 0.53)<br>p = 0.197 |
| Percentage of total drinks selected that are alcoholic<sup>4</sup> | Beta-binomial regression | -0.63, $p < 0.001$ | -1.27, $p < 0.001$ | -0.54, $p < 0.001$ |
| **Secondary outcomes: purchasing** |  |  |  |  |
|  |  | **Compared to reference group: 25% non-alcoholic (n = 145)** |  | **Reference group: 50% non-alcoholic (n = 141)** |
|  |  | **50% non-alcoholic (n = 141)** | **75% non-alcoholic (n = 136)** | **75% non-alcoholic (n = 136)\*** |
| Number of alcohol units purchased (including additional drinks from study categories only)<sup>3</sup> | Hurdle model part 1: binary outcomes | -0.27 (95% CI -1.22, 0.69)<br>$p = .584$ | -0.76 (95% CI -1.65, 0.13)<br>$p = .100$ | -0.49 (95% CI -1.33, 0.34)<br>$p = 0.248$ |
| | Hurdle model part 2: non-zero outcomes | 0.09 (95% CI -0.07, 0.25)<br>$p = 0.263$ | -0.16 (95% CI -0.32, 0.00)<br>$p = 0.056$ | -0.25 (95% CI -0.42, -0.09)<br>$p = 0.003$ |
| Number of alcohol units purchased (including all additional drinks)<sup>3</sup> | Hurdle model part 1: binary outcomes | 0.85 (95% CI 0.45, 1.25)<br>$p < 0.001$ | 1.30 (95% CI 0.89, 1.72)<br>$p < 0.001$ | 0.46 (95% CI 0.03, 0.88)<br>$p = 0.034$ |
| | Hurdle model part 2: non-zero outcomes | 0.06 (95% CI -0.11, 0.24)<br>$p = 0.471$ | -0.04 (95% CI -0.22, 0.14)<br>$p = 0.658$ | -0.10 (95% CI -0.28, 0.07)<br>$p = 0.255$ |
| Percentage of total drinks purchased that are alcoholic (including additional drinks from study categories only) <sup>4</sup> | Beta-binomial regression | -0.57, p = 0.003 | -1.09, p < 0.001 | -0.51, p = 0.004 |
| Percentage of total drinks purchased that are alcoholic (including all additional drinks) <sup>4</sup> | Beta-binomial regression | -0.42, p = 0.015 | -0.76, p < 0.001 | -0.33, p = 0.049 |

<sup>1</sup> Significance threshold is 0.0167 for a 5% alpha
<sup>2</sup>UK definition of alcohol units is used: in the US this is equivalent to 0.564 standard drinks
<sup>3</sup>Part 2 of the model is a negative binomial regression, therefore the back-transformed 95% confidence intervals become asymmetric. P values for hurdle models are based on z-statistics from a hurdle model fitted from the normal distribution using the glmmTMB routine in R [29]
<sup>4</sup>Beta binomial regression models used for analysis and p values were calculated using the “oad” R package for the analysis of over-dispersed data [48,49]

**Table 4.**
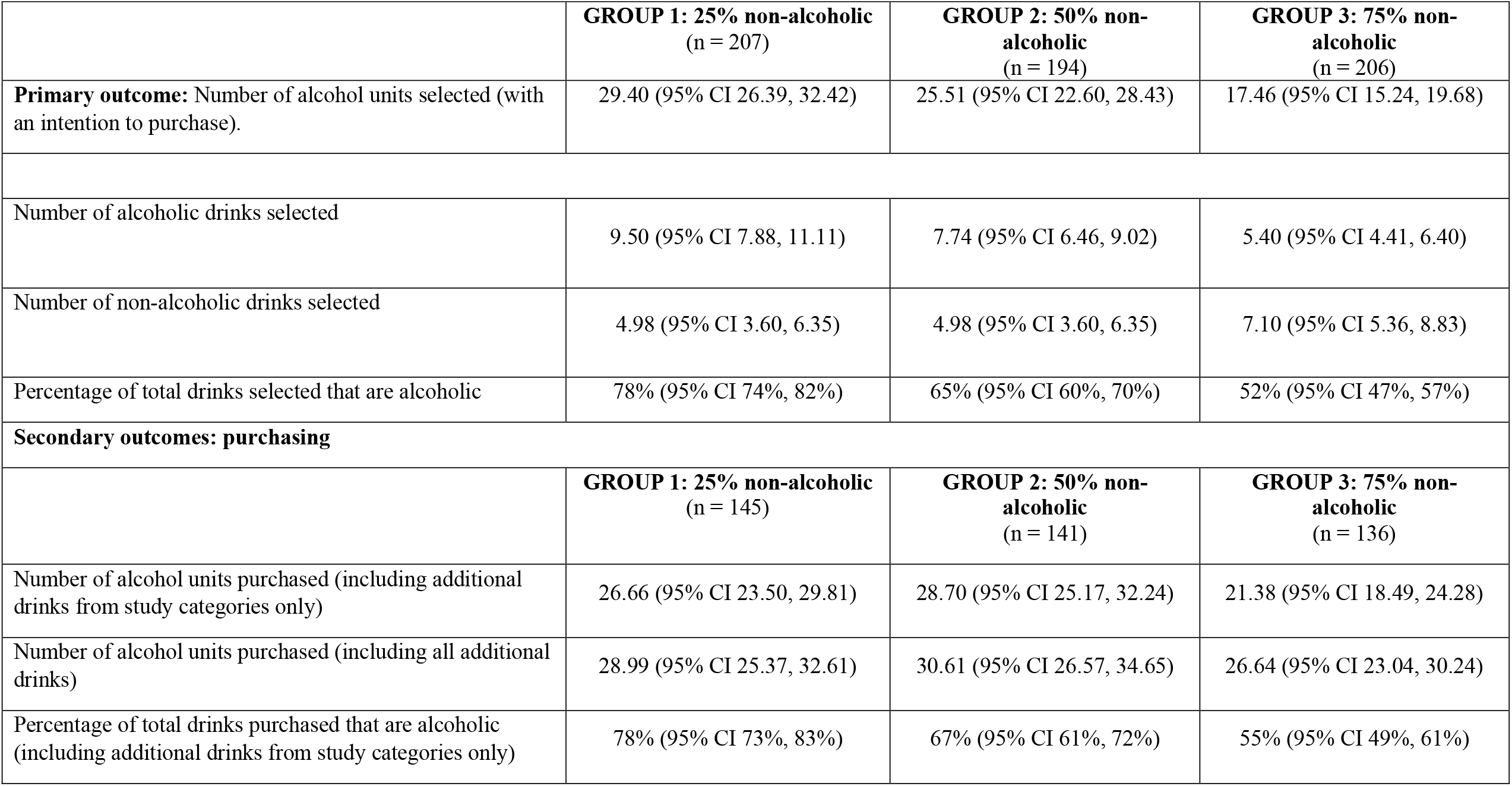
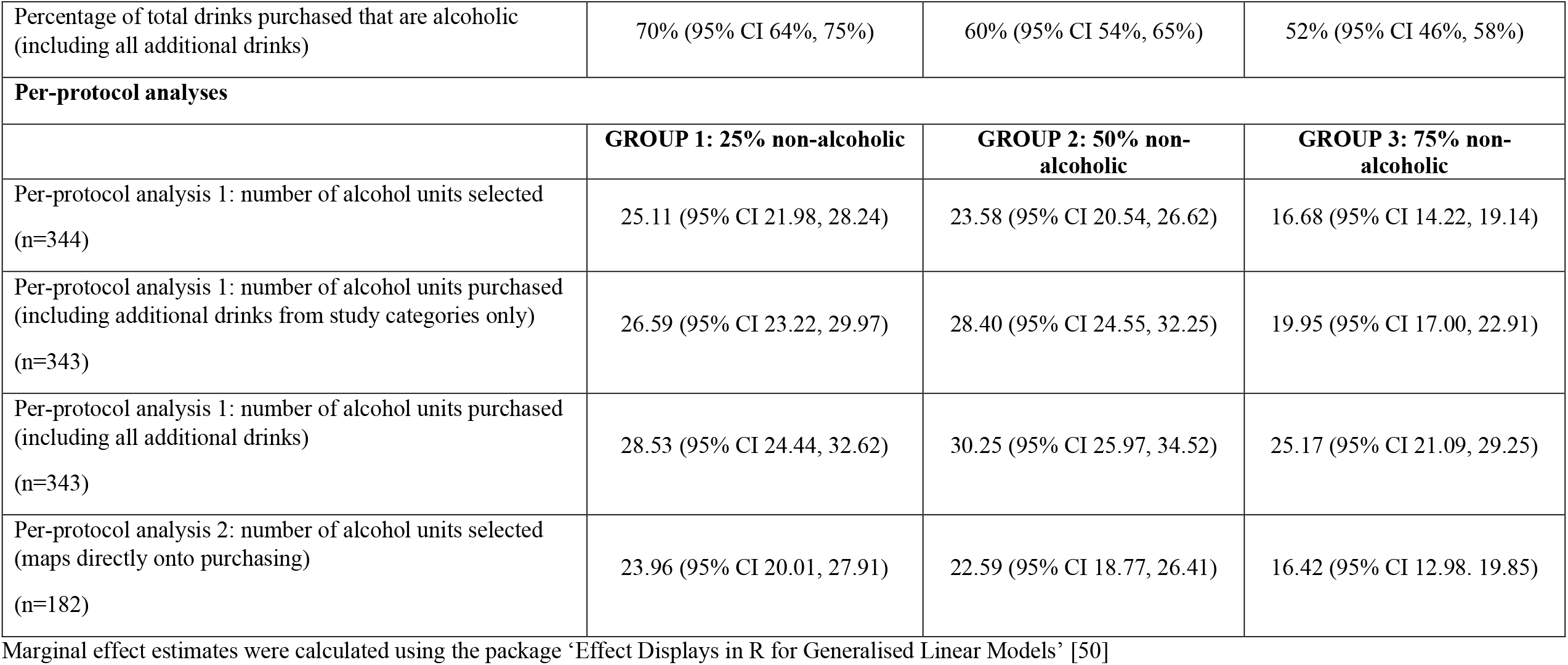
Marginal effect estimates predicted from hurdle models (95% confidence intervals) for primary and secondary outcomes.

|  | **GROUP 1: 25% non-alcoholic**<br>**(n = 207)** | **GROUP 2: 50% non-**<br>**alcoholic**<br>**(n = 194)** | **GROUP 3: 75% non-**<br>**alcoholic**<br>**(n = 206)** |
| --- | --- | --- | --- |
| **Primary outcome:** Number of alcohol units selected (with an intention to purchase). | 29.40 (95% CI 26.39, 32.42) | 25.51 (95% CI 22.60, 28.43) | 17.46 (95% CI 15.24, 19.68) |
| Number of alcoholic drinks selected | 9.50 (95% CI 7.88, 11.11) | 7.74 (95% CI 6.46, 9.02) | 5.40 (95% CI 4.41, 6.40) |
| Number of non-alcoholic drinks selected | 4.98 (95% CI 3.60, 6.35) | 4.98 (95% CI 3.60, 6.35) | 7.10 (95% CI 5.36, 8.83) |
| Percentage of total drinks selected that are alcoholic | 78% (95% CI 74%, 82%) | 65% (95% CI 60%, 70%) | 52% (95% CI 47%, 57%) |
| **Secondary outcomes: purchasing** |  |  |  |
|  | **GROUP 1: 25% non-alcoholic**<br>**(n = 145)** | **GROUP 2: 50% non-**<br>**alcoholic**<br>**(n = 141)** | **GROUP 3: 75% non-**<br>**alcoholic**<br>**(n = 136)** |
| Number of alcohol units purchased (including additional drinks from study categories only) | 26.66 (95% CI 23.50, 29.81) | 28.70 (95% CI 25.17, 32.24) | 21.38 (95% CI 18.49, 24.28) |
| Number of alcohol units purchased (including all additional drinks) | 28.99 (95% CI 25.37, 32.61) | 30.61 (95% CI 26.57, 34.65) | 26.64 (95% CI 23.04, 30.24) |
| Percentage of total drinks purchased that are alcoholic (including additional drinks from study categories only) | 78% (95% CI 73%, 83%) | 67% (95% CI 61%, 72%) | 55% (95% CI 49%, 61%) |
| Percentage of total drinks purchased that are alcoholic (including all additional drinks) | 70% (95% CI 64%, 75%) | 60% (95% CI 54%, 65%) | 52% (95% CI 46%, 58%) |
| **Per-protocol analyses** |  |  |  |
|  | **GROUP 1: 25% non-alcoholic** | **GROUP 2: 50% non-alcoholic** | **GROUP 3: 75% non-alcoholic** |
| Per-protocol analysis 1: number of alcohol units selected (n=344) | 25.11 (95% CI 21.98, 28.24) | 23.58 (95% CI 20.54, 26.62) | 16.68 (95% CI 14.22, 19.14) |
| Per-protocol analysis 1: number of alcohol units purchased (including additional drinks from study categories only) (n=343) | 26.59 (95% CI 23.22, 29.97) | 28.40 (95% CI 24.55, 32.25) | 19.95 (95% CI 17.00, 22.91) |
| Per-protocol analysis 1: number of alcohol units purchased (including all additional drinks) (n=343) | 28.53 (95% CI 24.44, 32.62) | 30.25 (95% CI 25.97, 34.52) | 25.17 (95% CI 21.09, 29.25) |
| Per-protocol analysis 2: number of alcohol units selected (maps directly onto purchasing) (n=182) | 23.96 (95% CI 20.01, 27.91) | 22.59 (95% CI 18.77, 26.41) | 16.42 (95% CI 12.98, 19.85) |

Marginal effect estimates were calculated using the package ‘Effect Displays in R for Generalised Linear Models’ [50]

### Primary outcome

Raw primary outcome data are presented in Table 2, modelled estimates for each part of the hurdle model in Table 3 and the overall marginal effect estimates in Table 4.

In the first part of the hurdle model, a greater proportion of participants in the ‘75% non-alcoholic’ group did not select any alcohol (27/206 [13.1%]) compared to the ‘25% non-alcoholic’ group (7/207 [3.4%]; 95% CI -2.09. -0.63; p < 0.001); there was no evidence of a difference between the ‘50% non-alcoholic’ (14/194 [7.2%]) and the ‘75% non-alcoholic’ group (95% CI 0.10, 1.34; p = 0.022, given the adjusted threshold of p = 0.0167) or between the ‘50% non-alcoholic group’ and the ‘25% non-alcoholic’ group (95% CI -1.44, 0.17; p = 0.121). In the second part of a hurdle model in participants (559/607) selecting any drinks containing alcohol, the ‘75% non-alcoholic’ group selected fewer alcohol units compared to the ‘50% non-alcoholic’ (95% CI -0.44, -0.14; p < 0.001) and ‘25% non-alcoholic’ (95% CI-0.54, -0.24; p < 0.001) groups, with no evidence of a difference between the ‘50% non-alcoholic’ and ‘25% non-alcoholic’ groups (95% CI -0.24, 0.05; p = 0.178). Overall, across all participants, 17.46 units (95% CI 15.24, 19.68) were selected in the ‘75% non-alcoholic’ group; 25.51 units (95% CI 22.60, 28.43) in the ‘50% non-alcoholic’ group; and 29.40 units (95% CI 26.39, 32.42) in the ‘25% non-alcoholic’ group. This corresponds to 8.1 fewer units (32% reduction) in the ‘75% non-alcoholic’ group compared to the ‘50% non-alcoholic’ group, and 11.9 fewer alcohol units compared to the ‘25% non-alcoholic’ group (a 41% reduction); 3.9 fewer units (13% reduction) were selected in the ‘50% non-alcoholic’ group than in the ‘25% non-alcoholic’ group.

### Secondary outcomes

Raw secondary outcome data are presented in Table 2, modelled estimates for each part of the hurdle model in Table 3 and the overall marginal effect estimates in Table 4. For purchasing outcomes, of the 640 participants who completed the selection task, 422 (66%) went on to purchase drinks from Tesco.com. Attrition from selection to purchasing stages was very similar across the three randomised groups (with 136, 141, and 145 completing purchasing).

Results for all secondary selection and purchasing outcomes demonstrated a wholly consistent pattern of results with amounts and proportions of alcohol selected and purchased consistently lowest in the ‘75% non-alcoholic’ group, although not always significantly so.

#### Selection

Participants in the ‘75% non-alcoholic’ group selected fewer alcoholic drinks than those in the ‘25% non-alcoholic’ group (95% CI -0.66, -0.20; p < 0.001), with an overall difference between marginal effect estimates of 4.1 drinks, equivalent to a 43% reduction. There were non-significant reductions in the ‘75% non-alcoholic’ compared to the ‘50% non-alcoholic’ group (95% CI -0.50, -0.02; p = 0.03; overall difference between marginal effect estimates of drinks, 30% reduction) and in the ‘50% non-alcoholic’ group compared to the ‘25% non-alcoholic’ group (95% CI -0.39, 0.06; p = 0.148; overall difference between marginal effect estimates of 1.8 drinks, 19% reduction).

There was no evidence of a difference in the number of non-alcoholic drinks selected between groups.

The percentage of total drinks selected that were alcoholic was lower in the ‘75% non-alcoholic’ group (52%, 95% CI 47%, 57%) compared to the ‘50% non-alcoholic’ group (65%, 95% CI 60%, 70%; p < 0.001), and lower compared to the ‘25% non-alcoholic’ group (78%, 95% CI 74%, 82%; p < 0.001); the percentage of total drinks selected that were alcoholic was also lower in the ‘50% non-alcoholic’ group compared to the ‘25% non-alcoholic’ group (p < 0.001).

#### Purchasing

When including additional drinks that were purchased from study categories only, there was a reduction in alcohol units purchased in the ‘75% non-alcoholic’ group compared to the ‘50% non-alcoholic’ group (95% CI -0.42, -0.09; p = 0.003, overall difference between marginal effect estimates of 7.3 drinks, 26% reduction). There was a non-significant reduction in the ‘75% non-alcoholic’ compared to the ‘25% non-alcoholic’ group (95% CI -0.32, -0.00; p = 0.056), with an overall difference between marginal effect estimates of 5.3 drinks, equivalent to a 20% reduction. There was no evidence of a difference in alcohol units purchased between the ‘50% non-alcoholic’ and the ‘25% non-alcoholic’ groups. There was evidence that the percentage of total drinks selected that were alcoholic was lower in the ‘75% non-alcoholic’ group (55%, 95% CI 49%, 61%) compared to the ‘50% non-alcoholic’ group (67%, 95% CI 61%, 72%; p = 0.004) and to the ‘25% non-alcoholic’ group (78%, 95% CI 73%, 83%; p < 0.001); the percentage of total drinks selected that were alcoholic was also lower in the ‘50% non-alcoholic’ group compared to the ‘25% non-alcoholic’ group (p = 0.003).

When including all additional drinks from any category, there was no evidence of a difference between any of the groups for alcohol units purchased. There was evidence of a difference in the percentage of total drinks selected that were alcoholic between the ‘75% non-alcoholic’ group (52%, 95% CI 46%, 58%) and the ‘25% non-alcoholic’ group (70%, 95% CI 64%, 75%; p < 0.001), and the ‘50% non-alcoholic’ group and the ‘25% non-alcoholic’ group (60%, 95% CI 54%, 65%; p = 0.015); there was no evidence of a difference between the ‘75% non-alcoholic’ and the ‘50% non-alcoholic’ groups.

### Per-protocol analyses

Of the 422 participants who purchased drinks, 344 participants purchased all the drinks they had selected in the selection task and 78 participants had one or more missing drinks. Of the 344 participants that purchased all the drinks they selected, 182 purchased no additional drinks.

Chi-squared tests indicated that there was no evidence against assuming equal attrition occurred. Exploratory analyses indicated that attrition was greater amongst participants with higher baseline alcohol purchasing, but regression using an interaction term suggested this did not bias the comparisons between groups, as there was no evidence of an effect at the usual threshold for interaction terms (p = 0.01) (Supplementary Material S4). See Table 5 for model results.

**Table 5.**
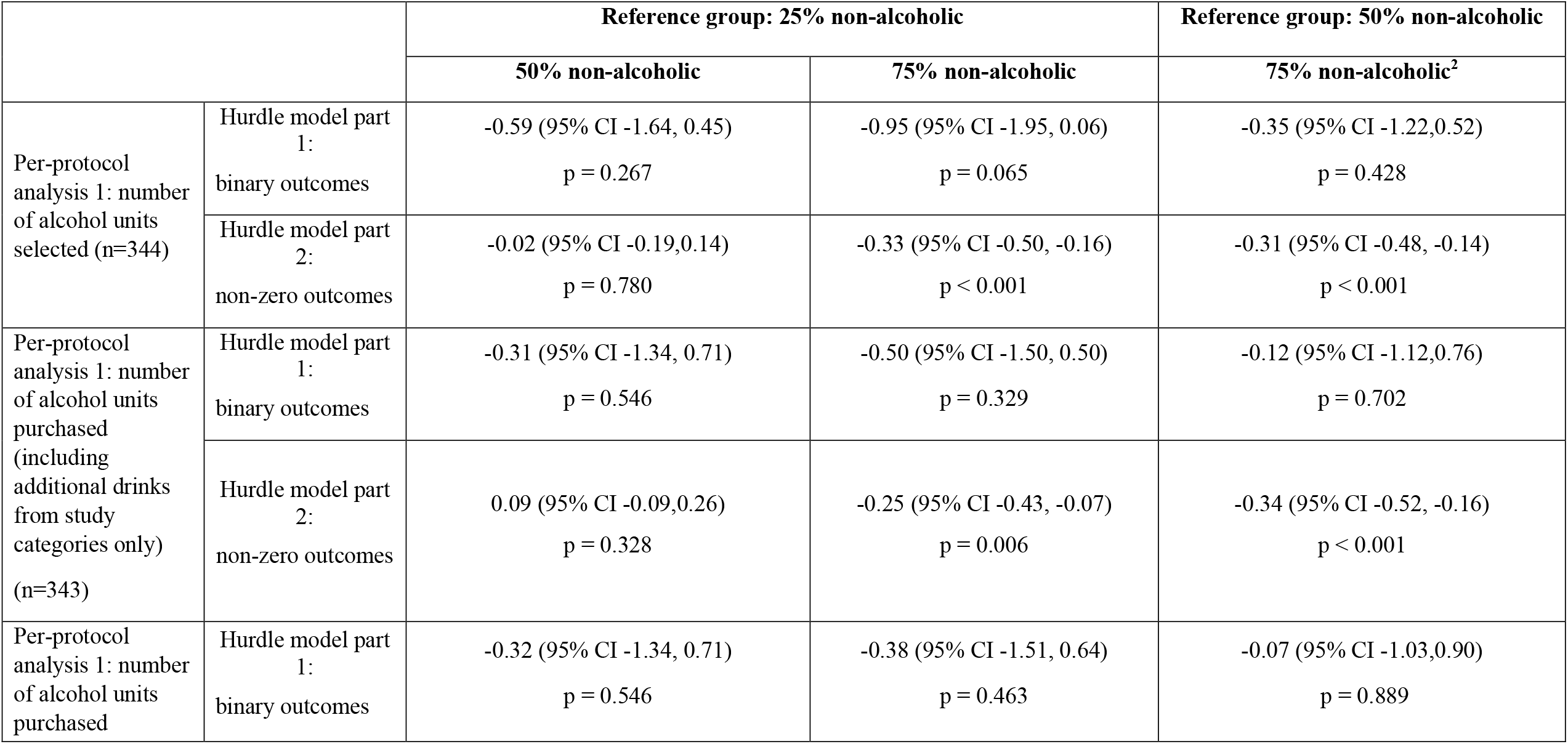
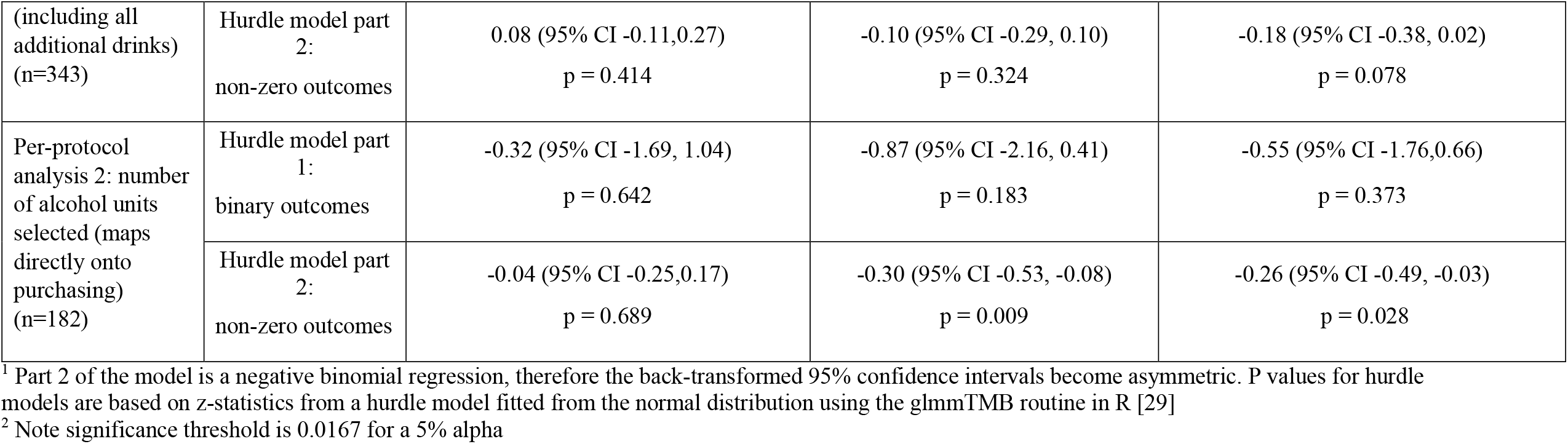
Per-protocol analyses for participants that purchased drinks: model estimates (95% confidence intervals), p values^1^.

In participants (n = 344) who completed purchasing of the drinks they had selected, either with or without additional drinks, those assigned to the ‘75% non-alcoholic’ group selected fewer alcohol units than the ‘50% non-alcoholic’ group (95% CI -0.48, -0.14; p < 0.001, overall difference between marginal effect estimates of 6.9 drinks, 29% reduction); the ‘75% non-alcoholic’ group also selected fewer alcohol units than those in the ‘25% non-alcoholic’ group (95% CI -0.50, -0.16; p < 0.001, overall difference between marginal effect estimates of 8.4 drinks, equivalent to a 34% reduction). There was no evidence of a difference between the ‘50% non-alcoholic’ and the ‘25% non-alcoholic’ groups. For purchasing, when including additional drinks from study categories only, fewer alcohol units were purchased in the ‘75% non-alcoholic’ group compared to the ‘50% non-alcoholic’ group (95% CI -0.52, -0.16; p < 0.001, overall difference between marginal effect estimates of 8.5 drinks, 30% reduction), and in the ‘75% non-alcoholic’ group compared to the ‘25% non-alcoholic’ group (95% CI - 0.43, -0.07; p = 0.006 overall difference between marginal effect estimates of 6.6 drinks, 25% reduction). There was no evidence of a difference between the ‘25% non-alcoholic’ and ‘50% non-alcoholic’ groups, and no evidence of a difference between groups for purchasing when including all additional drinks.

In participants (n = 182) who completed purchasing of only the drinks they had selected with no additional drinks, those assigned to the ‘75% non-alcoholic’ group selected and purchased fewer alcohol units than did those in the ‘25% non-alcoholic’ group (95% CI -0.53, -0.08; p = 0.009, overall difference between marginal effect estimates of 7.5 drinks, 31% reduction). There was no evidence of a difference for the other comparisons.

Full results for the additional outcomes can be found in Supplementary Material S5.

## Discussion

Our data show that substantially increasing the proportion of non-alcoholic drinks relative to alcoholic drinks meaningfully reduced the amount of alcohol selected and purchased in an online supermarket context. Compared to when the majority of options were alcoholic, participants selected 41% fewer alcohol units when the majority of options were non-alcoholic, and 32% fewer alcohol units when half the options were non-alcoholic.

Participants also went on to purchase significantly fewer alcohol units when the majority of options were non-alcoholic. Importantly, the overall pattern of results was consistent for all outcomes, with amounts and proportions of alcohol selected and purchased always lowest when non-alcoholic drinks were most available, including for pre-specified per-protocol analyses.

The findings of the current study are consistent with a single prior study that found increasing the proportion of non-alcoholic drinks options in an online setting reduced hypothetical selection of alcohol [12]. More generally, they are consistent with a growing body of studies that apply similar availability interventions to food [7,8,10], suggesting that these interventions have the potential to be usefully applied across different product contexts [5].

### Strengths and limitations

To our knowledge, this study is the first randomised controlled trial using a naturalistic setting to estimate the impact of increasing the proportion of non-alcoholic drinks. Meaningful selection and actual purchasing outcomes were assessed, with participants able to complete their typical online shop, including selecting and purchasing multiple options from a wide range of drinks.

The study had some limitations. First, while the primary selection outcome was assessed in the context of intention to subsequently purchase, and was minimally affected by attrition, there was substantial drop-out between selection and actual purchasing outcomes. However, attrition between groups was very similar by study condition, and there was sufficient power to detect effects despite this; as there is an absence of studies that look at purchasing of alcohol in this setting, effect sizes could not be anticipated, but large effects on purchasing were observed. While substantial attrition is expected in studies of this nature because of time between selection and purchasing, more generally it may be hard to avoid for any measure of unconstrained purchasing in a real-world online supermarket. Although we are not aware of other directly comparable studies in this context, more generally, ‘cart abandonment’ – where people do not purchase items they put in their shopping cart – is common in online (including supermarket) shopping contexts [31]. Future studies may be able address this through more intensive initial screening or follow-up of participants, or by forcing participants to immediately complete their online shop. However, such processes would arguably be less naturalistic, and including only the most motivated participants risks including a less representative sample.

Second, although the setting was as naturalistic as was feasible and actual purchasing outcomes were measured, the process involved two stages. Drinks were initially selected within a simulated online supermarket, before purchasing was completed in an actual online supermarket (albeit with the visual presentation of the former modelled on the latter). The principal purpose of including a measure of purchasing in the actual online supermarket was to validate and strengthen our primary outcome of selection, rather than to measure purchasing behaviour in a separate context. However, this meant that additional drink options were available in the real online supermarket, and participants could not be prevented from buying these if they wished to. As a result, the clearest effects on purchasing behaviour were in participants that followed the protocol as instructed and only purchased what they selected in the simulated supermarket where the intervention was implemented. To avoid this, the intervention would ideally have been implemented entirely within a real online supermarket. However, to our knowledge, this is the first study of an availability intervention to make use of such a setting (albeit in conjunction with a simulated supermarket component), although simulated retail settings, both online and physical, are commonly used in similar intervention studies [32-36]. This represents the most robust design used to date and could provide a useful method through which to assess interventions without requiring complex collaboration with commercial retailers, although further research is needed to assess its external validity. Finally, while participants were largely representative of Tesco.com shoppers [37] they were mostly of higher socioeconomic position. The generalisability of these findings to disadvantaged populations therefore needs consideration, particularly as buying alcohol-free drinks is more likely to occur in less socially and materially deprived households [18].

### Implications for research and policy

This study suggests that increasing the available non-alcoholic, and reducing the available alcoholic options has the potential to meaningfully reduce selection and purchasing of alcohol. Although there was some evidence of a reduction in alcohol selected and purchased when half of the options available were non-alcoholic, effects were only consistently observed when non-alcoholic drinks became the majority. Currently, supermarkets typically stock a wider range of alcoholic than non-alcoholic alternatives to alcohol, and these results suggest that if non-alcoholic options were to become the majority instead, we might expect to see substantial reductions in alcohol purchasing. As it is yet to be seen if such major changes in ranges of drinks are feasible in real-world settings, these findings are most reasonably interpreted as proof of principle, rather than able to directly inform policy options. It is plausible that this situation could rapidly change, however. The increase in popularity of alcohol-free drinks is relatively recent, with the global market growing substantially in the last four years, and in the UK it is forecast to continue to increase [15]. This recent increase in the popularity of alcohol-free drinks has led to the emergence of drinking settings reflecting this, such as an alcohol-free off licence in London [38]. In food retail contexts there have been substantial changes seen in healthier or more sustainable ranges - such as the introduction of 50% plant-based menus [39] and the requirement to provide at least 50% healthier options in healthcare settings in Scotland [40] - suggesting that shifts of such magnitude are possible. However, before policy recommendations are made, a robust evidence base suggesting potential effectiveness is required [5], which this study provides an initial step towards. Future studies should investigate the impact of smaller and more granular alterations in proportions of non-alcoholic drinks, and in a wider range of field settings, to establish how such interventions could be used. Given the relatively large effects observed in this study, subtler interventions could elicit smaller effects that would nonetheless remain meaningful for population health, especially when considering the inherent potential for scalability across retail settings.

This intervention simultaneously increased the number of non-alcoholic drinks and decreased the number of alcoholic drinks whilst the overall number of drinks remained constant. It is unclear whether the effect is predominantly driven by one or the combination of these changes. Further studies are needed to disentangle this and investigate potential mechanisms more broadly; noting that there is some preliminary exploration of possible mechanisms in food contexts [6,41,42]. Importantly, the overall number of drinks that participants selected and purchased remained similar between groups, suggesting that effects were a result of shifting, rather than necessarily restricting, choices. This implies overall drink sales and potentially revenues may be relatively unchanged if such an intervention were to be implemented, albeit dependent on non-alcoholic drink pricing. Increasing non-alcoholic drink availability could also ultimately lead to a greater range of alcohol-free drinks and soft drinks being manufactured, further increasing their popularity in synergistic fashion [18], and many alcohol companies have already committed to this [17]. It is important to note that because many alcohol-free alternatives are marketed by the alcohol industry and there is no regulation on the often-exaggerated health claims that are made about these drinks [43]. Such industry involvement has potential harms and should be monitored closely [44–47]. In addition, although some of the non-alcoholic drink options in the current study contained no sugar and were generally lower in calories than the alcoholic options (an average of 64 calories per non-alcoholic drink versus 233 calories per alcoholic drink), many soft drinks and alcohol-free alternatives still contain large amounts of sugar and calories. Given the health risks associated with sugary drink consumption [47], continued regulation and policies to reduce sugar content and consumption from both alcoholic and non-alcoholic drinks is needed to mitigate these risks.

## Conclusion

This randomised controlled trial is the first to date – to our knowledge – to assess the effect on selection and purchasing of increasing the proportion of non-alcoholic drinks available. The findings provide evidence that substantially increasing the proportion of non-alcoholic drinks - from 25% to 50% or 75% - meaningfully reduces alcohol selection and purchasing in an online supermarket context. While these findings highlight the potential for reducing alcohol sales at population level, further studies are warranted to assess whether these effects are realised in a range of real-world settings.

## Data Availability

Data will be available from the Open Science Framework (together with the study protocol and statistical analysis plan, already uploaded) and the University of Cambridge Research Repository upon publication.

https://osf.io/x726n/

Supplementary S1: Drink options and detailed task description

Supplementary S2. Number of zero values by group and outcome.

Supplementary S3. Secondary outcomes

Supplementary S4: Effect of attrition

Supplementary S5: Additional outcomes

## Declarations

### Authors’ contributions

GJH, NC, TMM, AKMB, and MRM conceptualized and designed the study. NC coordinated the study and led on data collection and cleaning with AKMB & JF. MAP and KDL led on the statistical analysis. NC, GJH, and MAP drafted the manuscript with all authors providing critical revisions. All authors had full access to all the data in the study and accept responsibility to submit for publication.

## Supplementary Material

**Supplementary S1: Drink options and detailed task description**

Alcohol-free beer, cider and wine were clearly labelled to ensure they were not confused with alcoholic drinks. Each drink option presented was a different brand, i.e., as the availability of non-alcoholic drink options increased, there was a larger choice of brands from which to choose. Drink images were all shown as bottles or cans, either as single items or multipacks, depending on the availability of products at Tesco online supermarket. Price promotions and variations for all drinks included in the selection task were checked every month via Tesco.com and recorded. Prices shown in the task therefore reflected the full price on Tesco.com for the duration of the study.

**Table S1a.** Proportions of non-alcoholic and alcoholic drinks displayed in the selection task.

| Drink range | Drink subset availability |  |  |
| --- | --- | --- | --- |
|  | Group 1: 25% non-alcoholic | Group 2: 50% non-alcoholic | Group 3: 75% non-alcoholic |
| Beer, cider and soft drinks (n) | <u>AF beer and cider (4)</u><br>AF lager (2)<br>AF ale (1)<br>AF cider (1)<br><u>Soft drinks (4)</u><br><u>Beer and cider (24)</u><br>Lager (10)<br>Ale (8)<br>Cider (6) | <u>AF beer and cider (8)</u><br>AF lager (3)<br>AF ale (3)<br>AF cider (2)<br><u>Soft drinks (8)</u><br><u>Beer and cider (16)</u><br>Lager (6)<br>Ale (6)<br>Cider (4) | <u>Alcohol-free (AF) beer and cider (12)</u><br>AF lager (4)<br>AF ale (4)<br>AF cider (4)<br><u>Soft drinks (12)</u><br><u>Beer and cider (8)</u><br>Lager (3)<br>Ale (3)<br>Cider (2) |
| Wine and soft drinks (n) | <u>AF wine (4)</u><br>AF red wine (1)<br>AF white wine (1)<br>AF rose or sparkling wine (2)<br><u>Soft drinks (4)</u><br><u>Wine (24)</u><br>Red wine (9)<br>White wine (9)<br>Rosé or sparkling wine (6) | <u>AF wine (8)</u><br>AF red wine (3)<br>AF white wine (3)<br>AF rose or sparkling wine (2)<br><u>Soft drinks (8)</u><br><u>Wine (16)</u><br>Red wine (6)<br>White wine (6)<br>Rosé or sparkling wine (4) | <u>AF wine (12)</u><br>AF red wine (3)<br>AF white wine (5)<br>AF rose or sparkling wine (4)<br><u>Soft drinks (12)</u><br><u>Wine (8)</u><br>Red wine (3)<br>White wine (3)<br>Rosé or sparkling wine (2) |

AF = Alcohol Free

**Table S1b.**
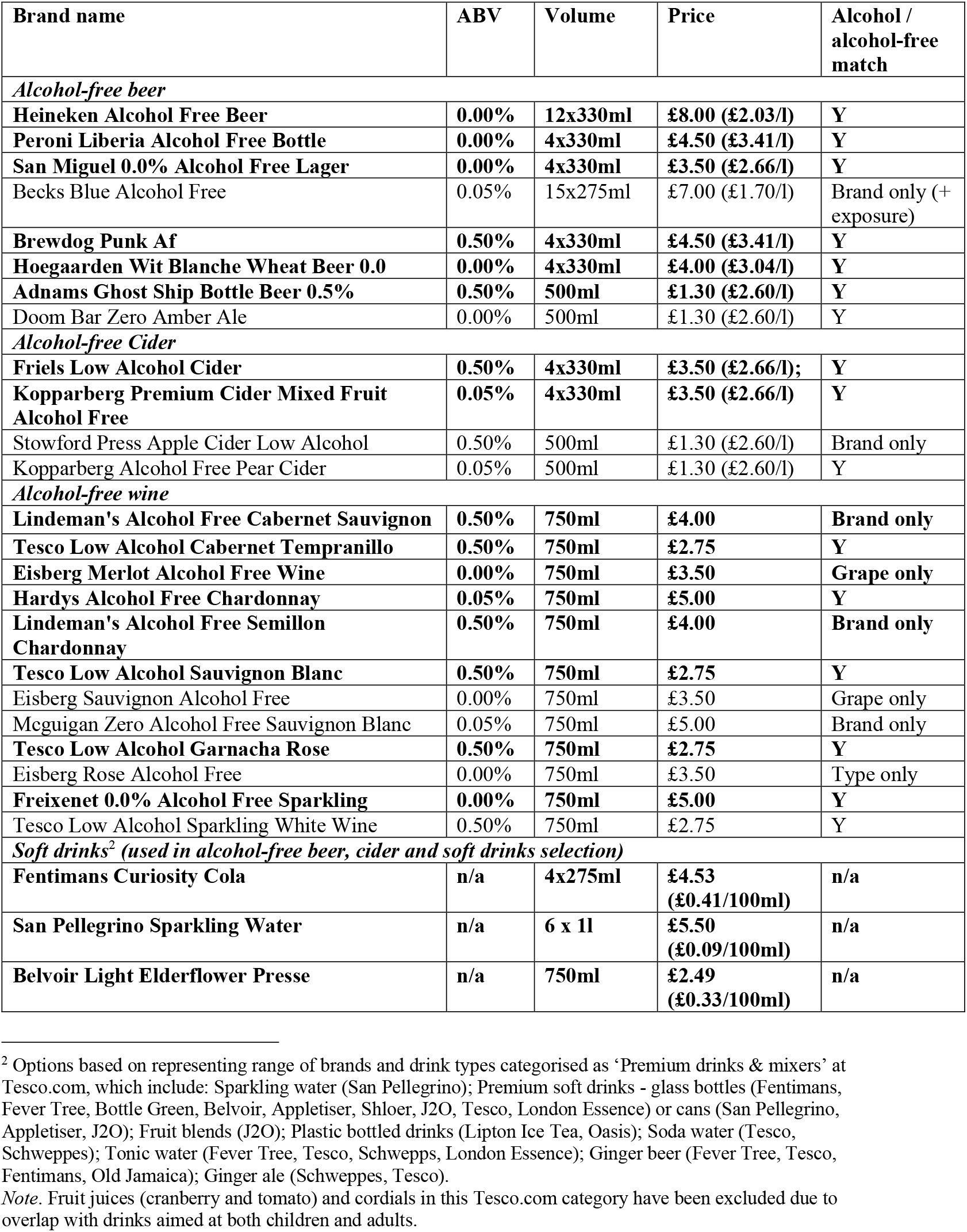
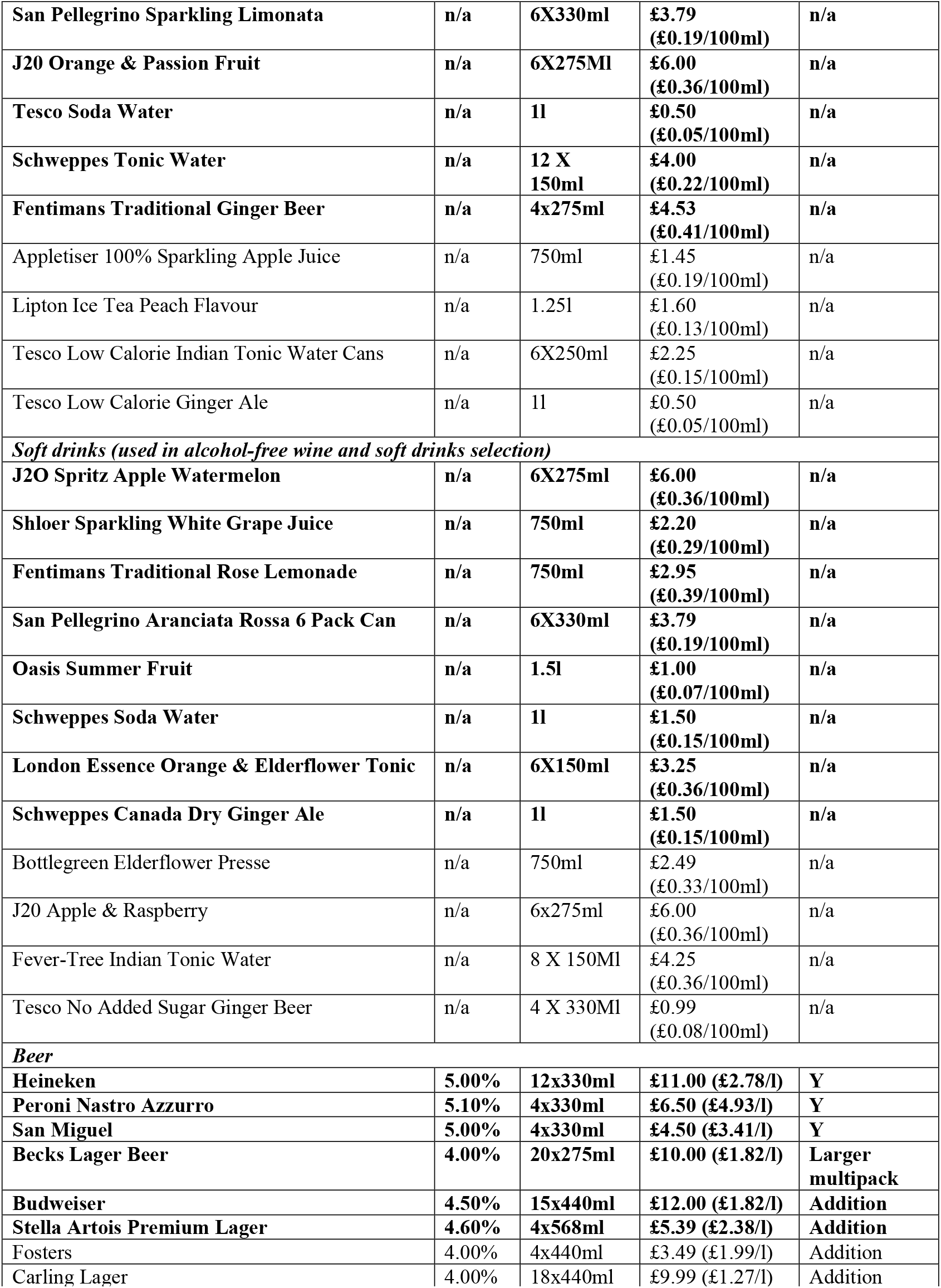
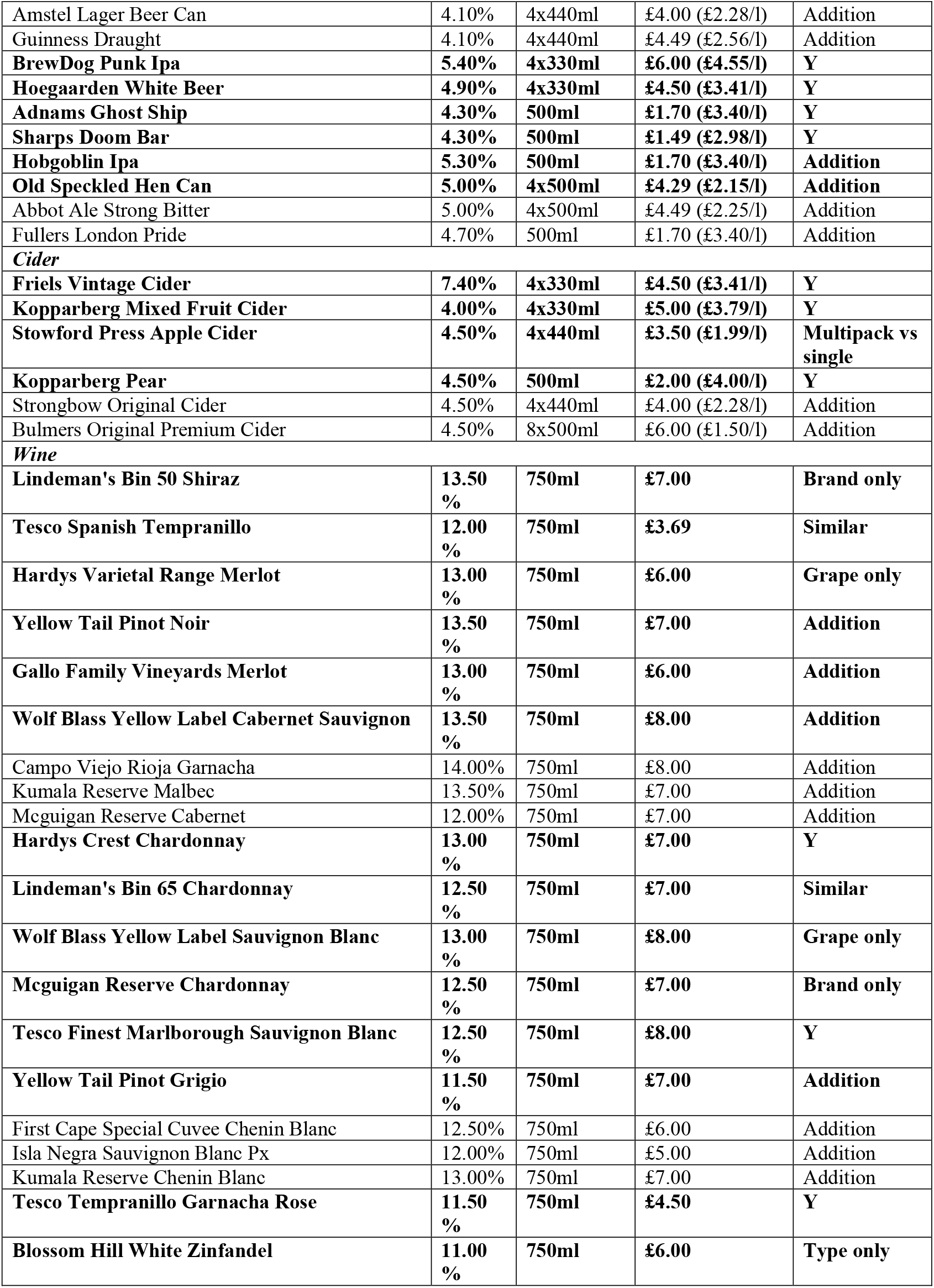
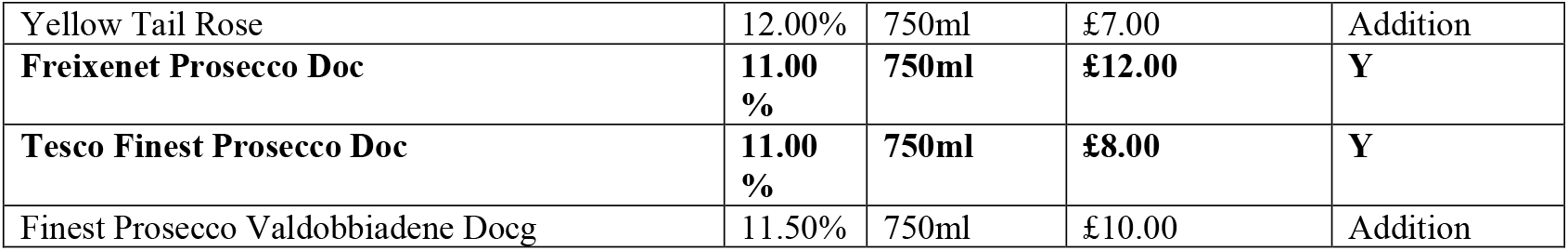
Drink options used in the selection task (prices based on Tesco.com, January 2021). Products shown in bold are those used in the 50% non-alcoholic condition.

| Brand name | ABV | Volume | Price | Alcohol / alcohol-free match |
| --- | --- | --- | --- | --- |
| *Alcohol-free beer* |  |  |  |  |
| **Heineken Alcohol Free Beer** | **0.00%** | **12x330ml** | **£8.00 (£2.03/l)** | **Y** |
| **Peroni Liberia Alcohol Free Bottle** | **0.00%** | **4x330ml** | **£4.50 (£3.41/l)** | **Y** |
| **San Miguel 0.0% Alcohol Free Lager** | **0.00%** | **4x330ml** | **£3.50 (£2.66/l)** | **Y** |
| Becks Blue Alcohol Free | 0.05% | 15x275ml | £7.00 (£1.70/l) | Brand only (+ exposure) |
| **Brewdog Punk Af** | **0.50%** | **4x330ml** | **£4.50 (£3.41/l)** | **Y** |
| **Hoegaarden Wit Blanche Wheat Beer 0.0** | **0.00%** | **4x330ml** | **£4.00 (£3.04/l)** | **Y** |
| **Adnams Ghost Ship Bottle Beer 0.5%** | **0.50%** | **500ml** | **£1.30 (£2.60/l)** | **Y** |
| Doom Bar Zero Amber Ale | 0.00% | 500ml | £1.30 (£2.60/l) | Y |
| *Alcohol-free Cider* |  |  |  |  |
| **Friels Low Alcohol Cider** | **0.50%** | **4x330ml** | **£3.50 (£2.66/l);** | **Y** |
| **Kopparberg Premium Cider Mixed Fruit Alcohol Free** | **0.05%** | **4x330ml** | **£3.50 (£2.66/l)** | **Y** |
| Stowford Press Apple Cider Low Alcohol | 0.50% | 500ml | £1.30 (£2.60/l) | Brand only |
| Kopparberg Alcohol Free Pear Cider | 0.05% | 500ml | £1.30 (£2.60/l) | Y |
| *Alcohol-free wine* |  |  |  |  |
| **Lindeman's Alcohol Free Cabernet Sauvignon** | **0.50%** | **750ml** | **£4.00** | **Brand only** |
| **Tesco Low Alcohol Cabernet Tempranillo** | **0.50%** | **750ml** | **£2.75** | **Y** |
| **Eisberg Merlot Alcohol Free Wine** | **0.00%** | **750ml** | **£3.50** | **Grape only** |
| **Hardys Alcohol Free Chardonnay** | **0.05%** | **750ml** | **£5.00** | **Y** |
| **Lindeman's Alcohol Free Semillon Chardonnay** | **0.50%** | **750ml** | **£4.00** | **Brand only** |
| **Tesco Low Alcohol Sauvignon Blanc** | **0.50%** | **750ml** | **£2.75** | **Y** |
| Eisberg Sauvignon Alcohol Free | 0.00% | 750ml | £3.50 | Grape only |
| Mcguigan Zero Alcohol Free Sauvignon Blanc | 0.05% | 750ml | £5.00 | Brand only |
| **Tesco Low Alcohol Garnacha Rose** | **0.50%** | **750ml** | **£2.75** | **Y** |
| Eisberg Rose Alcohol Free | 0.00% | 750ml | £3.50 | Type only |
| **Freixenet 0.0% Alcohol Free Sparkling** | **0.00%** | **750ml** | **£5.00** | **Y** |
| Tesco Low Alcohol Sparkling White Wine | 0.50% | 750ml | £2.75 | Y |
| *Soft drinks<sup>2</sup> (used in alcohol-free beer, cider and soft drinks selection)* |  |  |  |  |
| **Fentimans Curiosity Cola** | **n/a** | **4x275ml** | **£4.53 (£0.41/100ml)** | **n/a** |
| **San Pellegrino Sparkling Water** | **n/a** | **6 x 1l** | **£5.50 (£0.09/100ml)** | **n/a** |
| **Belvoir Light Elderflower Presse** | **n/a** | **750ml** | **£2.49 (£0.33/100ml)** | **n/a** |

<sup>2</sup> Options based on representing range of brands and drink types categorised as 'Premium drinks & mixers' at Tesco.com, which include: Sparkling water (San Pellegrino); Premium soft drinks - glass bottles (Fentimans, Fever Tree, Bottle Green, Belvoir, Appletiser, Shloer, J2O, Tesco, London Essence) or cans (San Pellegrino, Appletiser, J2O); Fruit blends (J2O); Plastic bottled drinks (Lipton Ice Tea, Oasis); Soda water (Tesco, Schweppes); Tonic water (Fever Tree, Tesco, Schweppes, London Essence); Ginger beer (Fever Tree, Tesco, Fentimans, Old Jamaica); Ginger ale (Schweppes, Tesco).
*Note.* Fruit juices (cranberry and tomato) and cordials in this Tesco.com category have been excluded due to overlap with drinks aimed at both children and adults.

|  |  |  |  |  |
| --- | --- | --- | --- | --- |
| San Pellegrino Sparkling Limonata | n/a | 6X330ml | £3.79<br>(£0.19/100ml) | n/a |
| J20 Orange & Passion Fruit | n/a | 6X275ml | £6.00<br>(£0.36/100ml) | n/a |
| Tesco Soda Water | n/a | 1l | £0.50<br>(£0.05/100ml) | n/a |
| Schweppes Tonic Water | n/a | 12 X<br>150ml | £4.00<br>(£0.22/100ml) | n/a |
| Fentimans Traditional Ginger Beer | n/a | 4x275ml | £4.53<br>(£0.41/100ml) | n/a |
| Appletiser 100% Sparkling Apple Juice | n/a | 750ml | £1.45<br>(£0.19/100ml) | n/a |
| Lipton Ice Tea Peach Flavour | n/a | 1.25l | £1.60<br>(£0.13/100ml) | n/a |
| Tesco Low Calorie Indian Tonic Water Cans | n/a | 6X250ml | £2.25<br>(£0.15/100ml) | n/a |
| Tesco Low Calorie Ginger Ale | n/a | 1l | £0.50<br>(£0.05/100ml) | n/a |
| *Soft drinks (used in alcohol-free wine and soft drinks selection)* |  |  |  |  |
| J20 Spritz Apple Watermelon | n/a | 6X275ml | £6.00<br>(£0.36/100ml) | n/a |
| Shloer Sparkling White Grape Juice | n/a | 750ml | £2.20<br>(£0.29/100ml) | n/a |
| Fentimans Traditional Rose Lemonade | n/a | 750ml | £2.95<br>(£0.39/100ml) | n/a |
| San Pellegrino Aranciata Rossa 6 Pack Can | n/a | 6X330ml | £3.79<br>(£0.19/100ml) | n/a |
| Oasis Summer Fruit | n/a | 1.5l | £1.00<br>(£0.07/100ml) | n/a |
| Schweppes Soda Water | n/a | 1l | £1.50<br>(£0.15/100ml) | n/a |
| London Essence Orange & Elderflower Tonic | n/a | 6X150ml | £3.25<br>(£0.36/100ml) | n/a |
| Schweppes Canada Dry Ginger Ale | n/a | 1l | £1.50<br>(£0.15/100ml) | n/a |
| Bottlegreen Elderflower Presse | n/a | 750ml | £2.49<br>(£0.33/100ml) | n/a |
| J20 Apple & Raspberry | n/a | 6x275ml | £6.00<br>(£0.36/100ml) | n/a |
| Fever-Tree Indian Tonic Water | n/a | 8 X 150ml | £4.25<br>(£0.36/100ml) | n/a |
| Tesco No Added Sugar Ginger Beer | n/a | 4 X 330ml | £0.99<br>(£0.08/100ml) | n/a |
| *Beer* |  |  |  |  |
| Heineken | 5.00% | 12x330ml | £11.00 (£2.78/l) | Y |
| Peroni Nastro Azzurro | 5.10% | 4x330ml | £6.50 (£4.93/l) | Y |
| San Miguel | 5.00% | 4x330ml | £4.50 (£3.41/l) | Y |
| Becks Lager Beer | 4.00% | 20x275ml | £10.00 (£1.82/l) | Larger<br>multipack |
| Budweiser | 4.50% | 15x440ml | £12.00 (£1.82/l) | Addition |
| Stella Artois Premium Lager | 4.60% | 4x568ml | £5.39 (£2.38/l) | Addition |
| Fosters | 4.00% | 4x440ml | £3.49 (£1.99/l) | Addition |
| Carling Lager | 4.00% | 18x440ml | £9.99 (£1.27/l) | Addition |
| Amstel Lager Beer Can | 4.10% | 4x440ml | £4.00 (£2.28/l) | Addition |
| Guinness Draught | 4.10% | 4x440ml | £4.49 (£2.56/l) | Addition |
| **BrewDog Punk Ipa** | **5.40%** | **4x330ml** | **£6.00 (£4.55/l)** | **Y** |
| **Hoegaarden White Beer** | **4.90%** | **4x330ml** | **£4.50 (£3.41/l)** | **Y** |
| **Adnams Ghost Ship** | **4.30%** | **500ml** | **£1.70 (£3.40/l)** | **Y** |
| **Sharps Doom Bar** | **4.30%** | **500ml** | **£1.49 (£2.98/l)** | **Y** |
| **Hobgoblin Ipa** | **5.30%** | **500ml** | **£1.70 (£3.40/l)** | **Addition** |
| **Old Speckled Hen Can** | **5.00%** | **4x500ml** | **£4.29 (£2.15/l)** | **Addition** |
| Abbot Ale Strong Bitter | 5.00% | 4x500ml | £4.49 (£2.25/l) | Addition |
| Fullers London Pride | 4.70% | 500ml | £1.70 (£3.40/l) | Addition |
| **Cider** |  |  |  |  |
| **Friels Vintage Cider** | **7.40%** | **4x330ml** | **£4.50 (£3.41/l)** | **Y** |
| **Kopparberg Mixed Fruit Cider** | **4.00%** | **4x330ml** | **£5.00 (£3.79/l)** | **Y** |
| **Stowford Press Apple Cider** | **4.50%** | **4x440ml** | **£3.50 (£1.99/l)** | **Multipack vs single** |
| **Kopparberg Pear** | **4.50%** | **500ml** | **£2.00 (£4.00/l)** | **Y** |
| Strongbow Original Cider | 4.50% | 4x440ml | £4.00 (£2.28/l) | Addition |
| Bulmers Original Premium Cider | 4.50% | 8x500ml | £6.00 (£1.50/l) | Addition |
| **Wine** |  |  |  |  |
| **Lindeman's Bin 50 Shiraz** | **13.50 %** | **750ml** | **£7.00** | **Brand only** |
| **Tesco Spanish Tempranillo** | **12.00 %** | **750ml** | **£3.69** | **Similar** |
| **Hardys Varietal Range Merlot** | **13.00 %** | **750ml** | **£6.00** | **Grape only** |
| **Yellow Tail Pinot Noir** | **13.50 %** | **750ml** | **£7.00** | **Addition** |
| **Gallo Family Vineyards Merlot** | **13.00 %** | **750ml** | **£6.00** | **Addition** |
| **Wolf Blass Yellow Label Cabernet Sauvignon** | **13.50 %** | **750ml** | **£8.00** | **Addition** |
| Campo Viejo Rioja Garnacha | 14.00% | 750ml | £8.00 | Addition |
| Kumala Reserve Malbec | 13.50% | 750ml | £7.00 | Addition |
| Mcguigan Reserve Cabernet | 12.00% | 750ml | £7.00 | Addition |
| **Hardys Crest Chardonnay** | **13.00 %** | **750ml** | **£7.00** | **Y** |
| **Lindeman's Bin 65 Chardonnay** | **12.50 %** | **750ml** | **£7.00** | **Similar** |
| **Wolf Blass Yellow Label Sauvignon Blanc** | **13.00 %** | **750ml** | **£8.00** | **Grape only** |
| **Mcguigan Reserve Chardonnay** | **12.50 %** | **750ml** | **£7.00** | **Brand only** |
| **Tesco Finest Marlborough Sauvignon Blanc** | **12.50 %** | **750ml** | **£8.00** | **Y** |
| **Yellow Tail Pinot Grigio** | **11.50 %** | **750ml** | **£7.00** | **Addition** |
| First Cape Special Cuvee Chenin Blanc | 12.50% | 750ml | £6.00 | Addition |
| Isla Negra Sauvignon Blanc Px | 12.00% | 750ml | £5.00 | Addition |
| Kumala Reserve Chenin Blanc | 13.00% | 750ml | £7.00 | Addition |
| **Tesco Tempranillo Garnacha Rose** | **11.50 %** | **750ml** | **£4.50** | **Y** |
| **Blossom Hill White Zinfandel** | **11.00 %** | **750ml** | **£6.00** | **Type only** |
| Yellow Tail Rose | 12.00% | 750ml | £7.00 | Addition |
| **Freixenet Prosecco Doc** | **11.00%** | **750ml** | **£12.00** | **Y** |
| **Tesco Finest Prosecco Doc** | **11.00%** | **750ml** | **£8.00** | **Y** |
| Finest Prosecco Valdobbiadene Docg | 11.50% | 750ml | £10.00 | Addition |

**Supplementary S2.** Number of zero values in the selection task by group and outcome.

| **Number of participants that:** | **Group 1: 25% non-alcoholic**<br>**(n = 207)** | **Group 2: 50% non-alcoholic**<br>**(n = 194)** | **Group 3: 75% non-alcoholic**<br>**(n = 206)** |
| --- | --- | --- | --- |
| Selected no drinks (alcoholic or non-alcoholic) | 2 | 4 | 5 |
| Selected no alcoholic drinks | 12 | 18 | 37 |
| Selected no non-alcoholic drinks | 120 | 72 | 56 |
| Selected no drinks containing alcohol units | 7 | 14 | 27 |

**Supplementary S3. Secondary outcomes**

A substantial number of datapoints met our pre-specified criterion of being possible outliers (Median Absolute Deviation (MAD) of >3) but these were typically considered to the part of the natural variability of such selection and purchasing measurements [1]. Due to concerns about unreasonable influence of any extreme or highly improbable values, we excluded one extreme value for units of alcohol purchased with a MAD of >15 (being more than two times the next highest value), with sales of over 290 units of alcohol (more than 50% more than the next highest value seen - 192.45 vs 291.85 - and relative to typical baseline purchasing and consumption by the same individual of only 0-2 units, and that worsened model fit.

**Supplementary S4: Effect of attrition**

**Table S4A:**
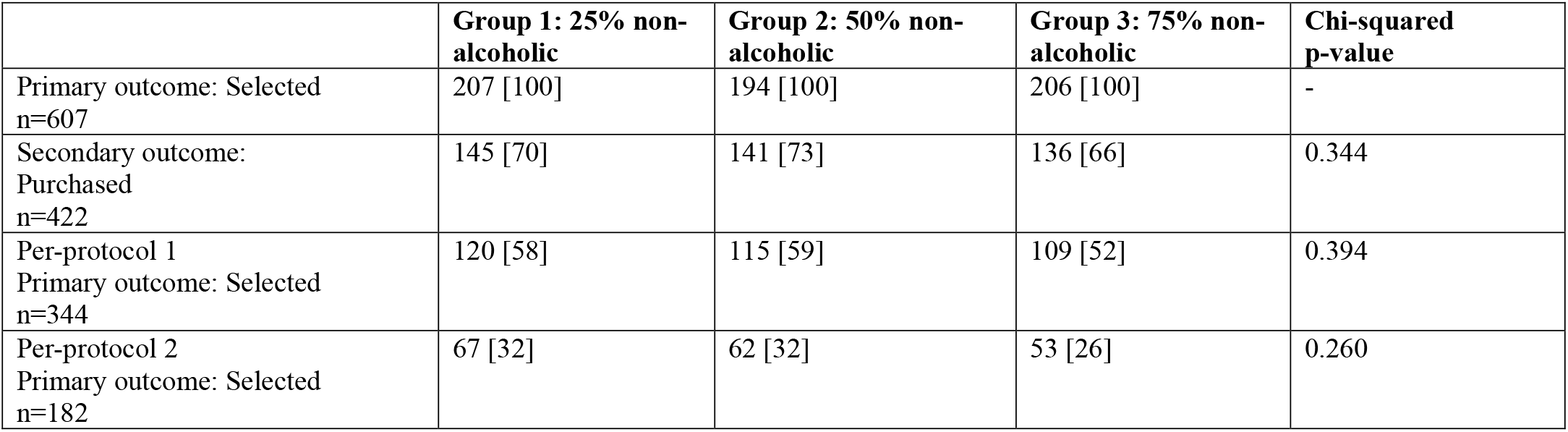
Effect of attrition from selection to purchasing on groups (n, [%])

|  | **Group 1: 25% non-alcoholic** | **Group 2: 50% non-alcoholic** | **Group 3: 75% non-alcoholic** | **Chi-squared p-value** |
| --- | --- | --- | --- | --- |
| Primary outcome: Selected<br>n=607 | 207 [100] | 194 [100] | 206 [100] | - |
| Secondary outcome: Purchased<br>n=422 | 145 [70] | 141 [73] | 136 [66] | 0.344 |
| Per-protocol 1<br>Primary outcome: Selected<br>n=344 | 120 [58] | 115 [59] | 109 [52] | 0.394 |
| Per-protocol 2<br>Primary outcome: Selected<br>n=182 | 67 [32] | 62 [32] | 53 [26] | 0.260 |

**Table S4B:** Effect of attrition from selection to purchasing on weekly units purchased at baseline (mean [median])

|  | **Group 1: 25% non-alcoholic** | **Group 2: 50% non-alcoholic** | **Group 3: 75% non-alcoholic** | **Model p-value\*: Attrition (interaction)** |
| --- | --- | --- | --- | --- |
| Primary outcome: Selected<br>n=607 | 40.6 [32.6] | 36.8 [29.3] | 40.9 [34.6] |  |
| Secondary outcome: Purchased<br>n=422 | 36.39 [32.6] | 37.5 [27.5] | 38.2 [31.9] | 0.125 (0.300) |
| Per-protocol 1<br>Primary outcome: Selected<br>n=344 | 34.7 [31.58] | 36.7 [27.5] | 36.7 [30.0] | 0.017 (0.396) |
| Per-protocol 2<br>Primary outcome: Selected<br>n=182 | 34.1 [28.0] | 34.7 [27.8] | 31.3 [27.2] | 0.024 (0.440) |

\*A model using sqrt (weekly units purchased) had acceptable regression diagnostics.

**Supplementary S5: Additional outcomes**

When including additional drinks from study categories only, those in the 75% non-alcoholic group purchased fewer alcoholic drinks than those in the 50% non-alcoholic group (-25%; 95%CIs -41%, - 6%; p = .015). There was no evidence of a difference between the 75% non-alcoholic and 25% non-alcoholic groups (-18%; 95%CIs -35%,4%; p = .103), or the 25% non-alcoholic and 50% non-alcoholic groups (10%; 95%CIs -12%,39%; p = .402). There was evidence that those in the 75% non-alcoholic group purchased more non-alcoholic drinks than those in the 50% non-alcoholic group (66%; 95%CIs 10%,149%; p = .014) and the 25% non-alcoholic group (78%; 95%CIs 18%,166%; p = .005). There was no evidence of a difference between the 25% non-alcoholic and 50% non-alcoholic groups (7%; 95%CIs -28%,60%; p = .734).

When including all additional drinks, there was no evidence of a difference between groups for alcoholic drinks purchased, although there were non-significant reductions between the 75% non-alcoholic and 25% non-alcoholic groups (-16%; 95%CIs -32%, 6%; p = .141), and the 75% non-alcoholic and Equal groups (-24%; 95%CIs -39%, -4%; p = .022). There was evidence of an increase in non-alcoholic drinks purchased between the 75% non-alcoholic and 25% non-alcoholic groups (60%; 95%CI 11%, 132%; p = .011), and non-significant increases between the 50% non-alcoholic and the 75% non-alcoholic groups (28%) and the 50% non-alcoholic and the 25% non-alcoholic groups (25%).

There was no evidence that the total number of drinks selected or purchased differed between groups.

**Table S5a.** Additional outcomes (raw means (SD))

|  | GROUP 1: 25% non-alcoholic | GROUP 2: 50% non-alcoholic | GROUP 3: 75% non-alcoholic |
| --- | --- | --- | --- |
|  | (n = 207) | (n = 194) | (n = 206) |
|  | Mean (SD) |  |  |
| Selection |  |  |  |
| Total number of drinks selected | 16.0 (21.4) | 15.2 (16.2) | 15.2 (18.8) |
| Secondary and additional outcomes – purchasing |  |  |  |
|  | GROUP 1: 25% non-alcoholic<br>(n = 145) | GROUP 2: 50% non-alcoholic<br>(n = 141) | GROUP 3: 75% non-alcoholic<br>(n = 136) |
| Total number of drinks purchased (including additional drinks from study categories only) | 13.27 (13.59) | 14.17 (13.65) | 14.71 (12.05) |
| Total number of drinks purchased (including all additional drinks) | 15.42 (14.62) | 17.45 (16.12) | 17.5 (15.21) |
| Number of alcoholic drinks purchased (including additional drinks from study categories only) | 9.06 (9.52) | 9.61 (11.61) | 7.23 (7.78) |
| Number of alcoholic drinks purchased (including all additional drinks) | 9.06 (9.52) | 9.61 (11.61) | 7.48 (7.86) |
| Number of non-alcoholic drinks purchased (including additional drinks from study categories only) | 4.21 (10.76) | 4.56 (6.0) | 7.49 (10.04) |
| Number of non-alcoholic drinks purchased (including all additional drinks) | 6.23 (11.96) | 7.72 (9.87) | 10.02 (13.52) |

**Table S5b.** Full model results for additional outcomes: model estimates (95%confidence interval), p values, percentage changes (95% confidence interval)

|  | Model used | Reference group: 25% non-alcoholic<br>(n = 207) |  | Reference group: 50% non-alcoholic<br>(n = 194) |
| --- | --- | --- | --- | --- |
|  |  | 50% non-alcoholic<br>(n = 194) | 75% non-alcoholic<br>(n = 207) | 75% non-alcoholic<br>(n = 207)* |
| Additional outcomes (selection): |  |  |  |  |
| Total number of drinks selected | Negative-binomial regression<br>(weekly units purchased included as covariate) | -0.01 (95%CI -0.20, 0.18)<br>p = 0.920<br>-1%<br>(95%CI -18%, 19%) | -0.04 (95%CI -0.22, 0.14)<br>p = 0.664<br>-4%<br>(95%CI -20%, 15%) | -0.03 (95%CI -0.22, 0.15)<br>p = 0.743<br>-3%<br>(95%CI -20%, 17%) |
| Additional outcomes (purchasing): |  |  |  |  |
| Total number of drinks purchased (including additional drinks from study categories only) | Negative-binomial regression | 0.08 (95%CI -0.11, 0.27)<br>p = 0.429<br>8%<br>(95%CI -11%, 31%) | 0.133 (95%CI -0.1, 0.33)<br>p = 0.175<br>14%<br>(95%CI -6%, 38%) | 0.06 (95%CI -0.14, 0.25)<br>p = 0.568<br>6%<br>(95%CI -13%, 28%) |
| Total number of drinks purchased (including all additional drinks) | Negative-binomial regression | 0.14 (95%CI -0.05, 0.33)<br>p = 0.141<br>15%<br>(95%CI -5%, 40%) | 0.15,= (95%CI -0.04, 0.35)<br>p = 0.118<br>17%<br>(95%CI -4%, 41%) | 0.01 (95%CI -0.18, 0.20)<br>p = 0.918<br>10%<br>(95%CI -17%, 23%) |
| Number of alcoholic drinks purchased (including additional drinks from study categories only) | Negative-binomial regression | 0.10 (95%CI -0.13, 0.33)<br>p = 0.409<br>10%<br>(95%CI -12%, 39%) | -0.20 (95%CI -0.43, 0.04)<br>p = 0.103<br>-18%<br>(95%CI -35%, 4%) | -0.29 (95%CI -0.52, -0.06)<br>p = 0.015<br>-25%<br>(95%CI -41%, -6%) |
| Number of alcoholic drinks purchased (including all additional drinks) | Negative-binomial regression | 0.10 (95%CI -0.13, 0.32)<br>p = 0.402<br>10%<br>(95%CI -12%, 38%) | -0.17 (95%CI -0.40, 0.06)<br>p = 0.141<br>-16%<br>(95%CI -32%, 6%) | -0.27 (95%CI -0.50, -0.04)<br>p = 0.022<br>-24%<br>(95%CI -39%, -4%) |
| Number of non-alcoholic drinks purchased (including additional drinks from study categories only) | Negative-binomial regression | 0.07 (95%CI -0.33, 0.47)<br>p = 0.734<br>7%<br>(95%CI -28%, 60%) | 0.57 (95%CI 0.17, 0.98)<br>p = 0.005<br>78%<br>(95%CI 18%, 166%) | 0.50 (95%CI 0.10, 0.91)<br>p = 0.014<br>66%<br>(95%CI 10%, 149%) |
| Number of non-alcoholic drinks purchased (including all additional drinks) | Negative-binomial regression | 0.23 (95%CI -0.14, 0.59)<br>p = 0.223<br>25%<br>(95%CI -13%, 80%) | 0.47 (95%CI 0.10, 0.84)<br>p = 0.011<br>60%<br>(95%CI 11%, 132%) | 0.25 (95%CI -0.12, 0.62)<br>p = 0.187<br>28%<br>(95%CI -11%, 85%) |

<sup>1</sup>Note significance threshold is 0.0167 for a 5% alpha

## References

1. Murray CJL, Aravkin AY, Zheng P, Abbafati C, Abbas KM, Abbasi-Kangevari M, et al. Global burden of 87 risk factors in 204 countries and territories, 1990–2019: a systematic analysis for the Global Burden of Disease Study 2019. Lancet. 2020;396: 1223–1249.

2. World Health Organisation [WHO]. Global status report on alcohol and health; 2018 [cited April 2020]. Available from http://www.who.int/substance_abuse/publications/global_alcohol_report/en/

3. Hollands GJ, Bignardi G, Johnston M, Kelly MP, Ogilvie D, Petticrew M, et al. The TIPPME intervention typology for changing environments to change behaviour. Nat Hum Behav. 2017;1: 0140.

4. Marteau TM, White M, Rutter H, Petticrew M, Mytton OT, McGowan JG, et al. Increasing healthy life expectancy equitably in England by 5 years by 2035: could it be achieved? Lancet. 2019;393: 2571–2573.

5. Marteau TM, Hollands GJ, Pechey R, Reynolds JP, Jebb SA. Changing the assortment of available food and drink for leaner, greener diets. BMJ. 2022;377: e069848.

6. Pechey R, Hollands GJ, Carter P, Marteau TM. Altering the availability of products within physical micro-environments: a conceptual framework. BMC Public Health. 2020;20: 986.

7. Hollands GJ, Carter P, Anwer S, King SE, Jebb SA, Ogilvie D, et al. Altering the availability or proximity of food, alcohol, and tobacco products to change their selection and consumption. Cochrane Database Syst Rev. 2019; Issue 9: CD012573.

8. Reynolds JP, Ventsel M, Kosite D, Dames BR, Brocklebank L, Masterton S, et al. Impact of decreasing the proportion of higher energy foods and reducing portion sizes on food purchased in worksite cafeterias: A stepped-wedge randomised controlled trial. PLoS Med. 2021;18: e1003743.

9. Pechey R, Bateman P, Cook B, Jebb SA. Impact of increasing the relative availability of meat-free options on food selection: two natural field experiments and an online randomised trial. Int J Behav Nutr Phys Act. 2022;19: 9.

10. Garnett EE, Balmford A, Sandbrook C, Pilling MA, Marteau TM. Impact of increasing vegetarian availability on meal selection and sales in cafeterias. National Academy of Sciences. 2019;116: 20923–20929.

11. Brimblecombe J, McMahon E, Ferguson M, Silva KD, Peeters A, Miles E, et al. Effect of restricted retail merchandising of discretionary food and beverages on population diet: a pragmatic randomised controlled trial. Lancet Planet Health. 2020;4: e463–e473.

12. Blackwell AKM, De-loyde K, Hollands GJ, Morris RW, Brocklebank LA, Maynard OM, et al. The impact on selection of non-alcoholic vs alcoholic drink availability: an online experiment. BMC Public Health. 2020;20: 526. doi:10.1186/s12889-020-08633-5

13. The Morning Advertiser. The boom of alcohol-free is a sticking trend; 2019. [cited April 2020]. Available from https://www.morningadvertiser.co.uk/Article/2019/05/20/How-much-has-the-no-alcohol-category-grow

14. Financial Times. UK sales of low-alcohol and no-alcohol beers almost double in 5 years; 2022 [cited Oct 2022]. Available from https://www.ft.com/content/e7ff0844-06f3-4b6a-89d8-ea2f2a329cff

15. Statistica. Non-Alcoholic Beer - United Kingdom | Statista Market Forecast. 2022 [cited Sep 2022]. Available from https://www.statista.com/outlook/cmo/alcoholic-drinks/beer/non-alcoholic-beer/united-kingdom

16. IWSR. No- and Low-Alcohol in Key Global Markets Reaches Almost US$10 Billion in Value. 2022. [cited Feb 2022] Available from https://www.theiwsr.com/no-and-low-alcohol-in-key-global-markets-reaches-almost-us10-billion-in-value/

17. Department of Health and Social Care [DHSC]. Advancing our health: prevention in the 2020s – consultation document 2021. [cited Dec 2021]. Available from https://www.gov.uk/government/consultations/advancing-our-health-prevention-in-the-2020s/advancing-our-health-prevention-in-the-2020s-consultation-document

18. Anderson P, O’Donnell A, Kokole D, Jané Llopis E, Kaner E. Is Buying and Drinking Zero and Low Alcohol Beer a Higher Socio-Economic Phenomenon? Analysis of British Survey Data, 2015–2018 and Household Purchase Data 2015–2020. Int J Environ Res Public Health. 2021;18: 10347

19. Anderson P, Jané Llopis E, O’Donnell A, Manthey J, Rehm J. Impact of low and no alcohol beers on purchases of alcohol: interrupted time series analysis of British household shopping data, 2015–2018. BMJ Open. 2020;10: e036371.

20. Rehm J, Lachenmeier DW, Llopis EJ, Imtiaz S, Anderson P. Evidence of reducing ethanol content in beverages to reduce harmful use of alcohol. Lancet Gastroenterol Hepatol. 2016;1:78–83.

21. Statistica. Leading brands of beer in United Kingdom 2020. 2020 [cited Feb 2022]. Available from https://www.statista.com/statistics/868499/leading-brands-of-beer-in-the-uk/

22. Statistica. Leading brands of cider in the UK 2020. 2020 [cited Feb 2022]. Available from https://www.statista.com/statistics/317609/leading-brands-of-cider-in-the-uk/

23. Statistica. Leading brands of still wine in the UK 2020. 2020 [cited Feb 2022]. Available: from https://www.statista.com/statistics/304150/leading-brands-of-wine-including-sparkling-gb-in-the-uk/

24. Pechey R, Hollands GJ, Reynolds JP, Jebb SA, Marteau TM. Is altering the availability of healthier vs. less-healthy options effective across socioeconomic groups? A mega-analysis. Int J Behav Nutr Phys Act. 2022;19: 88.

25. Cleavebooks. Equivalent Unit Drinks Calculator. 2022 [cited Aug 2022]. Available from http://www.cleavebooks.co.uk/scol/ccalcoh1.htm

26. ONS. Primary set of harmonised concepts and questions [cited Feb 2022]. Available from https://webarchive.nationalarchives.gov.uk/ukgwa/20160106185646/ http://www.ons.gov.uk/ons/guide-method/harmonisation/primary-set-of-harmonised-concepts-and-questions/index.html

27. NI Direct. Qualifications: what the different levels mean; 2015. [cited Feb 2022]. Available from https://www.nidirect.gov.uk/articles/qualifications-what-different-levels-mean

28. Saunders JB, Aasland OG, Babor TF, de la Fuente JR, Grant M. Development of the Alcohol Use Disorders Identification Test (AUDIT): WHO Collaborative Project on Early Detection of Persons with Harmful Alcohol Consumption--II. Addiction 1993; 88: 791–804

29. Brooks ME, Kristensen K, Benthem KJ van, Magnusson A, Berg CW, Nielsen A, et al. glmmTMB Balances Speed and Flexibility Among Packages for Zero-inflated Generalized Linear Mixed Modeling. R Journal. 2017;9: 378–400.

30. Zeileis A, Kleiber C, Jackman S. Regression Models for Count Data in R. J Stat Softw. 2008;27: 1–25.

31. Shopping cart abandonment rate by industry 2021. In: Statista. [cited Dec 2021]. Available: https://www.statista.com/statistics/457078/category-cart-abandonment-rate-worldwide/

32. Hartmann-Boyce J, Bianchi F, Piernas C, Payne Riches S, Frie K, Nourse R, et al. Grocery store interventions to change food purchasing behaviors: a systematic review of randomized controlled trials. Am J Clin Nutr. 2018;107: 1004–1016.

33. Payne Riches S, Aveyard P, Piernas C, Rayner M, Jebb SA. Optimising swaps to reduce the salt content of food purchases in a virtual online supermarket: A randomised controlled trial. Appetite. 2019;133: 378–386.

34. Clarke N, Blackwell AKM, De-loyde K, Pechey E, Hobson A, Pilling M, et al. Health warning labels and alcohol selection: a randomised controlled experiment in a naturalistic shopping laboratory. Addiction. 2021;116: 3333–3345.

35. Clarke N, Blackwell AK, Ferrar J, De-loyde K, Pilling MA, Munafo MR, et al. Impact on alcohol selection and purchasing of increasing the proportion of non-alcoholic versus alcoholic drinks: randomised controlled trial. medRxiv; 2022. p. 2022.03.04.22271898.

36. Hall MG, Grummon AH, Higgins ICA, Lazard AJ, Prestemon CE, Avendaño-Galdamez MI, et al. The impact of pictorial health warnings on purchases of sugary drinks for children: A randomized controlled trial. PLoS Med. 2022;19: e1003885.

37. IDG. Trading with Tesco. 2017 [cited Feb 2022]. Available: <>https://shoppervista.igd.com/articles/article-viewer/t/infographic---trading-with-tesco/i/16633>

38. Club Soda. Club Soda opens London’s first alcohol-free off-licence. In: Club Soda. 2021 [cited Feb 2022]. Available: https://joinclubsoda.com/first-alcohol-free-off-licence

39. Vegconomist. Wagamama Confirms 50% of Menu Will Be Plant-Based By October - vegconomist: the vegan business magazine. 2021 [cited Feb 2022]. Available: <>https://vegconomist.com/gastronomy-and-catering/wagamama-confirms-50-of-menu-will-be-plant-based-by-october/>

40. http://Gov.uk. Criteria for the Healthcare Retail Standard. 2016 [cited Aug 2022]. Available: http://www.gov.scot/publications/criteria-healthcare-retail-standard-1/

41. Pechey R, Clarke N, Pechey E, Ventsel M, Hollands GJ, Marteau T. Impact of altering the available food options on selection: potential mediation by social norms. Appetite 2021;105245

42. Pechey R, Hollands GJ, Marteau T. Explaining the effect on food selection of altering availability: two experimental studies on the role of prior preferences. BMC Public Health 2022;22: 868.

43. Corfe S, Hyde R, Shepherd J. Alcohol-free and low-strength drinks. Social Market Foundation. 2020;95. Available from https://www.smf.co.uk/publications/no-low-alcohol-harms/

44. Mialon M, McCambridge J. Alcohol industry corporate social responsibility initiatives and harmful drinking: a systematic review. Eur J Public Health. 2018;28: 664–673

45. Petticrew M, Maani N, Pettigrew L, Rutter H, Van Schalkwyk MC. Dark Nudges and Sludge in Big Alcohol: Behavioral Economics, Cognitive Biases, and Alcohol Industry Corporate Social Responsibility. Milbank Q. 2020;98: 1290–1328.

46. Alcohol Health Alliance. NoLo product placement in soap operas: Alcohol health promotion or alibi branding? In: Alcohol Health Alliance. 2022 [cited May 2022]. Available from https://ahauk.org/nolo-product-placement-in-soap-operas-alcohol-health-promotion-or-alibi-branding/

47. Malik VS, Hu FB. The role of sugar-sweetened beverages in the global epidemics of obesity and chronic diseases. Nat Rev Endocrinol. 2022;18: 205–218.

48. Crowder MJ. Beta-Binomial Anova for Proportions. J R Stat Soc Ser C Appl Stat. 1978;27: 34–37.

49. Prentice RL. Binary Regression Using an Extended Beta-Binomial Distribution, With Discussion of Correlation Induced by Covariate Measurement Errors. J Am Stat Assoc. 1986;81: 321–327.

50. Fox J. Effect Displays in R for Generalised Linear Models. J Stat Softw. 2003;8: 1–27.

## References

1. Reynolds JP, Ventsel M, Kosīte D, Dames BR, Brocklebank L, Masterton S, et al. Impact of decreasing the proportion of higher energy foods and reducing portion sizes on food purchased in worksite cafeterias: A stepped-wedge randomised controlled trial. PLoS Med. 2021;18: e1003743.

